# A Phase 2 Randomized, Double-Blind, Placebo-controlled Trial of Oral Camostat Mesylate for Early Treatment of COVID-19 Outpatients Showed Shorter Illness Course and Attenuation of Loss of Smell and Taste

**DOI:** 10.1101/2022.01.28.22270035

**Authors:** Geoffrey Chupp, Anne Spichler-Moffarah, Ole S. Søgaard, Denise Esserman, James Dziura, Lisa Danzig, Reetika Chaurasia, Kailash P. Patra, Aryeh Salovey, Angela Nunez, Jeanine May, Lauren Astorino, Amisha Patel, Stephanie Halene, Jianhui Wang, Pei Hui, Prashant Patel., Jing Lu, Fangyong Li, Geliang Gan, Stephen Parziale, Lily Katsovich, Gary V. Desir, Joseph M. Vinetz

## Abstract

**Importance:** Early treatment of mild SARS-CoV-2 infection might lower the risk of clinical deterioration in COVID-19.

**Objective:** To determine whether oral camostat mesylate would reduce upper respiratory SARS-CoV-2 viral load in newly diagnosed outpatients with mild COVID-19, and would lead to improvement in COVID-19 symptoms.

**Design:** From June, 2020 to April, 2021, we conducted a randomized, double-blind, placebo-controlled phase 2 trial.

**Setting:** Single site, academic medical center, outpatient setting in Connecticut, USA.

**Participants:** Of 568 COVID-19 positive potential adult participants diagnosed within 3 days of study entry and assessed for eligibility, 70 were randomized and 498 were excluded (198 did not meet eligibility criteria, 37 were not interested, 265 were excluded for unknown or other reasons). The primary inclusion criteria were a positive SARS-CoV-2 nucleic acid amplification result in adults within 3 days of screening regardless of COVID-19 symptoms.

**Intervention:** Treatment was 7 days of oral camostat mesylate, 200 mg po four times a day, or placebo.

**Main Outcomes and Measures:** The primary outcome was reduction of 4-day log_10_ nasopharyngeal swab viral load by 0.5 log_10_ compared to placebo. The main prespecified secondary outcome was reduction in symptom scores as measured by a quantitative Likert scale instrument, Flu-PRO-Plus modified to measure changes in smell/taste measured using FLU-PRO-Plus.

**Results:** Participants receiving camostat had statistically significant lower quantitative symptom scores (FLU-Pro-Plus) at day 6, accelerated overall symptom resolution and notably improved taste/smell, and fatigue beginning at onset of intervention in the camostat mesylate group compared to placebo. Intention-to-treat analysis demonstrated that camostat mesylate was not associated with a reduction in 4-day log_10_ NP viral load compared to placebo.

**Conclusions and relevance:** The camostat group had more rapid resolution of COVID-19 symptoms and amelioration of the loss of taste and smell. Camostat compared to placebo was not associated with reduction in nasopharyngeal SARS-COV-2 viral load. Additional clinical trials are warranted to validate the role of camostat mesylate on SARS-CoV-2 infection in the treatment of mild COVID-19.

**Trial registration: Clinicaltrials.gov, NCT04353284 (04/20/20):** **(https://clinicaltrials.gov/ct2/show/NCT04353284?term=camostat+%2C+yale&draw=2&rank=1)**

**Key Points:** *Question:* Will early treatment of COVID-19 with a repurposed medication, camostat mesylate, improve clinical outcomes?

*Findings:* In this phase 2 randomized, double-blind placebo-controlled clinical trial that included 70 adults with early COVID-19, the oral administration of camostat mesylate treatment within 3 days of diagnosis prevented the loss of smell/taste and reduced the duration of illness.

*Meaning:* In the current COVID-19 pandemic, phase III testing of an inexpensive, repurposed drug for early COVID-19 is warranted.

---

Early treatment of COVID-19 has the potential to prevent clinical deterioration in the course of of SARS-CoV-2 infection. As the pandemic continues, data to support the use of efficacious oral treatments for outpatients with COVID-19 have not yet been published (although direct antiviral agents have been reported from Merck and Pfizer by press release) although parenterally-administered monoclonal antibody treatment is available through Emergency Use Authorization (EUA).^1^

SARS-CoV-2 uses the angiotensin converting enzyme II (ACE2) as its cell entry receptor to infect human cells.^2^ This mechanism exploits the human epithelial cell (respiratory, gastrointestinal tracts) surface-expressed Transmembrane Serine Protease 2 (TMPRSS2).^2-4^ Camostat mesylate, a protease inhibitor that inhibits the activity of TMPRSS2, is approved in Japan for the treatment of postoperative reflux esophagitis and acute exacerbations of chronic pancreatitis,^5^ is well tolerated and has no known drug-drug interactions. Previous studies have demonstrated that camostat mesylate blocks SARS-CoV-2 cellular entry *in vitro*.^2-4^ Camostat and structurally-related, parenterally-administered nafamostat improved disease pathogenesis and survival in mouse models of SARS-CoV-1^6^ and SARS-CoV-2,^7^ respectively.

This trial tested the hypothesis that camostat mesylate given to outpatients with SARS-CoV-2 infection will reduce upper respiratory viral burden. Pre-specified secondary outcomes were selected to determine whether there might be beneficial clinical effects of camostat mesylate.

## METHODS

This phase 2 clinical trial was designed early in the COVID-19 pandemic before detailed biology of the infection was known. The rationale for the duration of treatment with camostat mesylate (200 mg PO qid daily for 7 days) was in part based on the known viral kinetics of SARS-CoV-2 in COVID-19 patients, as well as the treatment guidelines for other lower respiratory tract viral infections. Because participants in this trial were enrolled early after outpatient diagnosis, we anticipated that most would present with mild to moderate disease. Therefore, the treatment principles for other lower respiratory tract infections of similar disease severity were followed in this protocol.

## TRIAL DESIGN AND OVERSIGHT

The trial was investigator-initiated and conducted at a single site (Yale New Haven Health System). Ono Pharmaceuticals (Osaka, Japan) provided study drug (FOIPAN) but did not influence study design or analysis, and did not provide other financial support. An independent data safety monitoring board (DSMB) oversaw the study. The study pharmacy prepared study drug by over-encapsulation with an identically prepared placebo. This was a prospective, single center, 1:1 randomized, placebo-controlled, double-blind Phase 2 clinical trial with eligible patients randomly assigned to receive either camostat mesylate or placebo for 7 days. Details of diagnostic testing and viral load quantification, randomization, blinding, trial oversight, and the collaboration model for the trial are provided in the Supplementary Appendix.

Seventy of the proposed 114 subjects were randomized before the DSMB recommended study closure. The prespecified interim analysis was conducted under the auspices of the DSMB with data prepared by the unblinded statisticians. The authors and the sponsors vouch for the completeness and accuracy of the data and for the fidelity of the trial to the protocol (available at NEJM.org).

The major inclusion criteria were age 18 or older, a first positive COVID-19 RT-PCR assay within the 3 days before enrollment, with at least one COVID-19-compatible symptom such as fever, upper respiratory symptoms, cough, chills, loss of taste/smell (see COVID-19-PRO symptom score sheet), or a recent high-risk exposure to COVID-19. Major exclusion criteria were hospitalized patients with Covid 19, pregnancy, lactation or an allergy to camostat mesylate. Detailed inclusion and exclusion criteria are available in the study protocol. No participants had been vaccinated for COVID-19 or had received monoclonal antibody therapy either before or during the trial.

The analyses were conducted and the manuscript was written by the first two and last authors and the trial statisticians. All the authors contributed to subsequent drafts, and no others contributed to the writing. The trial was conducted in accordance with the amended Declaration of Helsinki, and the protocol was approved by Yale’s Institutional Review Board (IRB). All the patients provided written informed consent.

This trial was carried out in accordance with the International Conference on Harmonization Good Clinical Practice (ICH GCP) and authroized by the U.S. Food and Drug Administration as a Phase 2 Investigational New Drug (IND) Trial. The Principal Investigator ensured that no deviation from or changes to the protocol took place without prior agreement from the IND sponsor, funding agency, and documented approval from the IRB, except where necessary to eliminate an immediate hazard to the trial participants.

## ASSESSMENT OF ADVERSE EVENTS

Adverse events were assessed by verbal report from the patient to the study coordinator, and were specifically sought according to the approval package insert. All adverse events had their relationship to the study intervention assessed by the clinician who examined and evaluated the participant based on a temporal relationship and his/her clinical judgment. The degree of certainty about causality was graded as related or not related.

## STATISTICAL DESIGN

### Sample size calculation

The sample size calculation was based on the primary endpoint, a change in the log_10_ respiratory (NP swab RT-PCR) viral load from baseline to day 4 post-randomization. Given the limited data on the variability of the change in log_10_ viral load, we powered the study based on detecting a moderate standardized effect size of 0.3 using an analysis of covariance (ANCOVA), adjusting for baseline log_10_ viral load. With a power of 90%, and a type I error rate of 10% (2-sided), we would be able to detect the hypothesized 0.3 standardized effect size with 98 patients divided into 49 patients per group with a 1:1 randomization. We increased the target sample size by 15% (i.e. n=114, 57 per treatment group), 5% for an efficacy and futility look at 50% information (i.e., when half of the patients were enrolled) and 10% to account for loss to follow up.

### Data analysis

The primary analysis was according to intent-to-treat. All analyses were performed using SAS 9.4 (Cary, NC).

### Primary Outcome

The primary endpoint was change in the log_10_ viral load of a NP swab specimen as determined by quantitative RT-PCR testing from baseline to day 4 post-randomization. To make use of all available data the treatment difference was estimated using a repeated measures linear mixed model analysis. As viral load decreases to zero over time, only timepoints up to day 6 (i.e., day 0, 2, 4, 6) were included to assure a sufficient number of non-zero levels. This model fits the same mean to both treatment groups at baseline (day 0) and thus conditions (i.e., adjusts) the estimates of treatment effects on baseline viral load.^8^ The t-statistic for the contrast of the treatment means at day 4 was used to determine whether camostat was statistically different from the placebo at the overall 10% level of significance.

An interim analysis for efficacy and futility using a group sequential method with asymmetric boundaries was conducted when 57 participants accumulated the primary endpoint. The early stopping boundary for efficacy was a t-statistic for the treatment difference greater than the critical value of 1.875. The boundary for futility was -1, indicating that camostat was trending in the wrong direction. Because of the interim analysis, a p-value of 0.06 (2-sided), corresponding to an efficacy boundary value of 1.875 at the last look, was used for the level of significance for the primary outcome at the final analysis.

### Predefined Secondary Outcomes

Because the clinical and virological natural history of COVID-19 infection was unclear at the outset of this trial, a series of secondary outcomes were pre-specified: change in log_10_ NP swab and saliva viral load from baseline to days 2 and 6; rate of positive tests at days 6, 14, and 28; and detailed symptom score quantification using a modified FLU-PRO instrument used for influenza,^9^ designated FLU-PRO-Plus (Supplementary Information), in which an additional question regarding smell and taste was added.

All statistical tests for secondary outcomes were conducted at 5% level of significance (two-sided) with no adjustment for multiple testing. Changes in viral load at days 2 and 6 (NP and saliva) as well as FLU-PRO symptom scores at days 6 and 14 were compared using similar mixed models as for the primary outcome. Proportions of positive COVID-19 tests at days 6, 14, and 28 were compared using Fisher’s exact tests. A repeated measures mixed model with a negative binomial distribution was used to compare loss of smell and taste between groups.

## RESULTS

### PATIENTS

Of 568 potential subjects assessed for eligibility (Fig. 1) from June 2020 to April 2021, 70 were randomized to receive camostat mesylate or placebo and analyzed in an intent-to-treat analysis; the largest group (n=198) was excluded for not meeting inclusion criteria. Baseline demographic features were generally similar between the two groups,including baseline viral loads determined by RT-PCR of the N, S and ORF1ab genes by NP swab, saliva samples and the FLU-PRO-plus symptom scores, which reflected mild illness. The placebo group had a higher proportion of subjects with at least one co-morbidity (97.1%) compared to the camostat mesylate group (82.9%) (Table 1). The mean (+/-SD) time from symptom onset until enrollment was not prospectively determined but was generally in the 2-3 day range.

**Figure 1.**
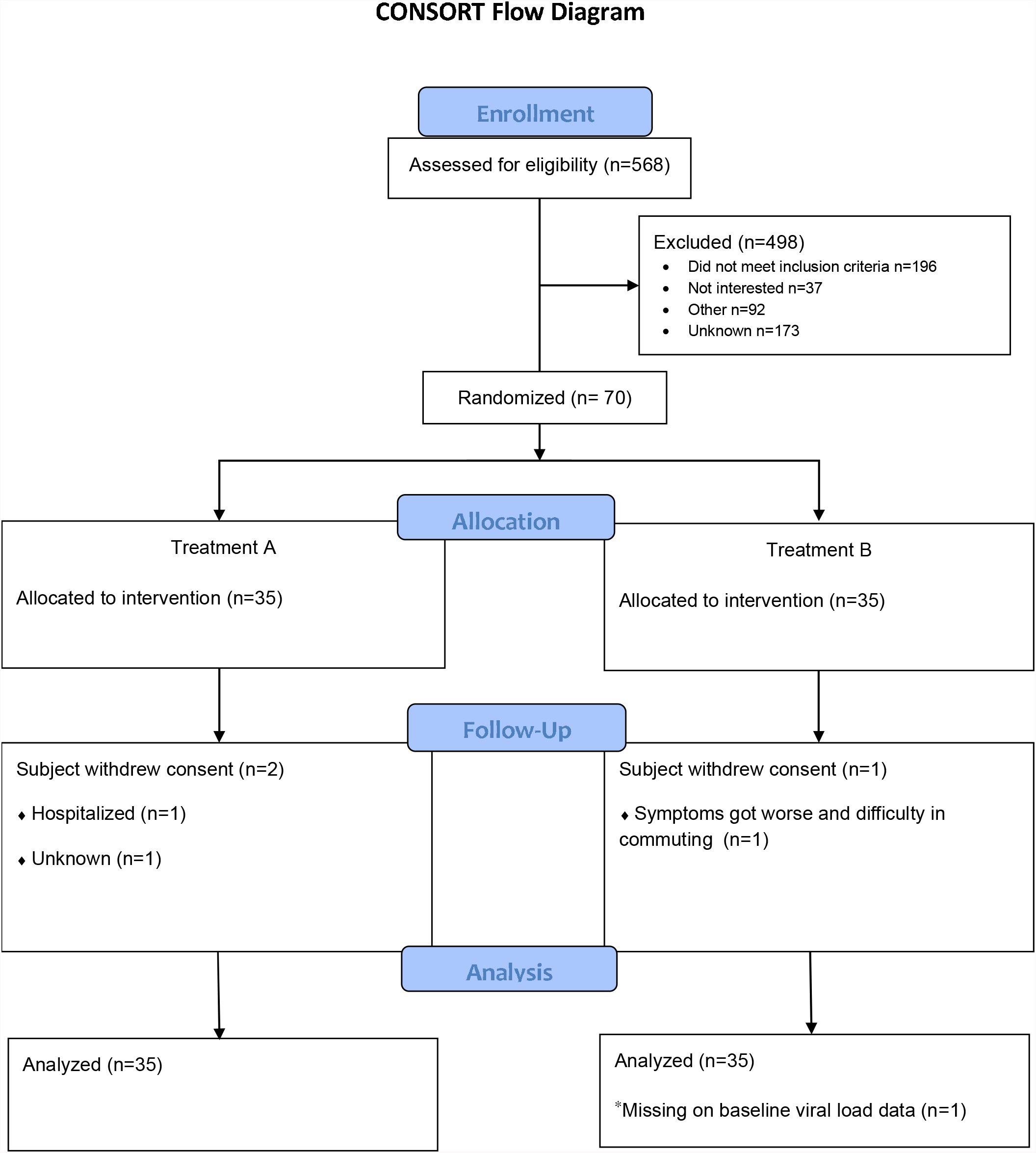
Enrollment, Randomization, and Analysis Populations

On April 22, 2021 the DSMB reviewed the pre-planned interim analysis of 57 patients with primary endpoint results and recommended that the study cease new enrollment of participants and complete follow-up and data collection on those enrolled. This decision was based on the crossing of a futility boundary for the primary outcome along with the new availability of outpatient treatment options including monoclonal antibody treatments. Ultimately 70 subjects were enrolled.

### VIRAL LOAD

The viral load in the camostat group was not significantly different at day 4 compared to placebo (Figure 2). The camostat group tended to have lower 4-day reductions in log_10_ NP viral loads compared to placebo (LSMEAN change= -2.0 camostat, -2.8 placebo, difference in means (95% CI) = 0.74 (−0.03, 1.51), p=0.06). In addition, there were no significant difference in NP swab nor saliva viral load between treatment groups at days 2 and 6 (Figure 2).

**Figure 2.**
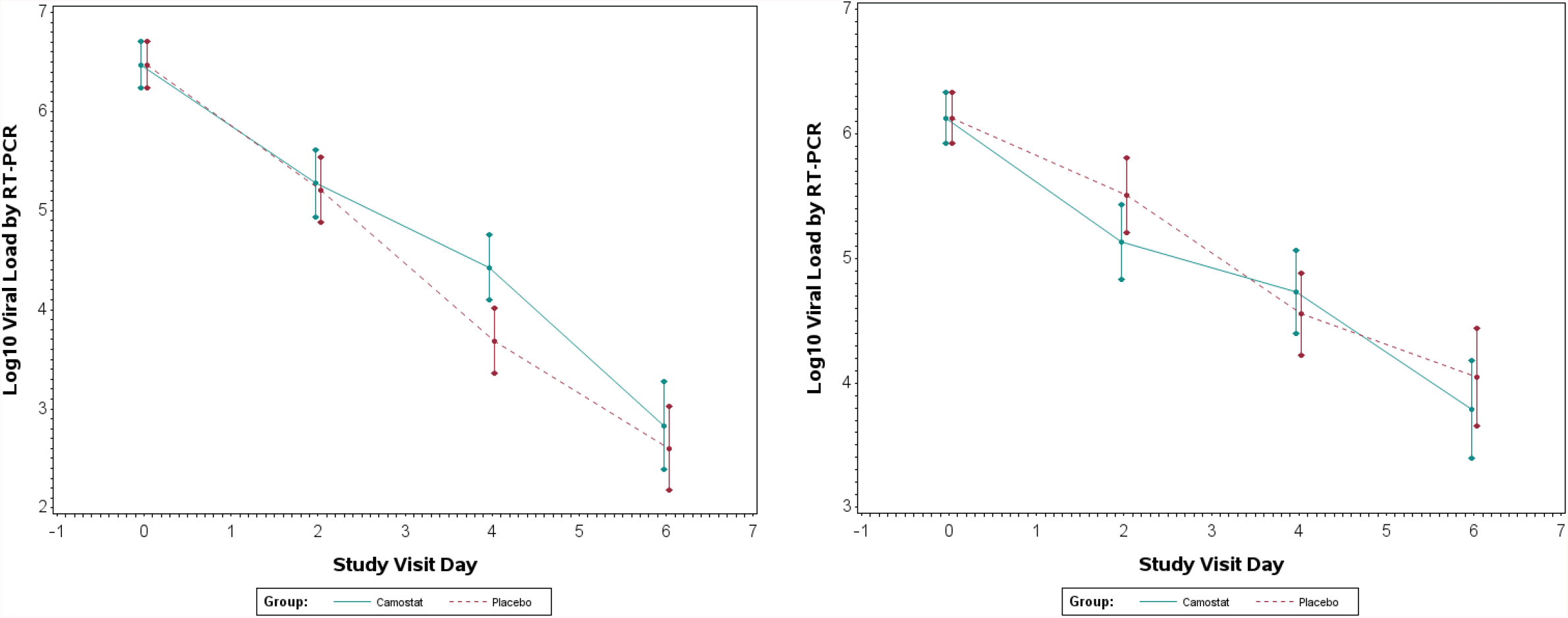
Comparison of nasopharyngeal swab and saliva viral loads in camostat vs. placebo groups.

### SYMPTOMS

Symptom severity scores at day 2 were similar between groups (Figure 3). However, greater reductions in scores at day 6 were observed in camostat (−19.1) compared to placebo (−12.5) (difference in means (95% CI) = -6.6 (−12.1, -1.2), p=0.02). Exploratory analyses revealed that camostat participants had lower scores for loss of taste and smell at days 2, 3, 4 and 5 (Figure 3).

**Figure 3.**
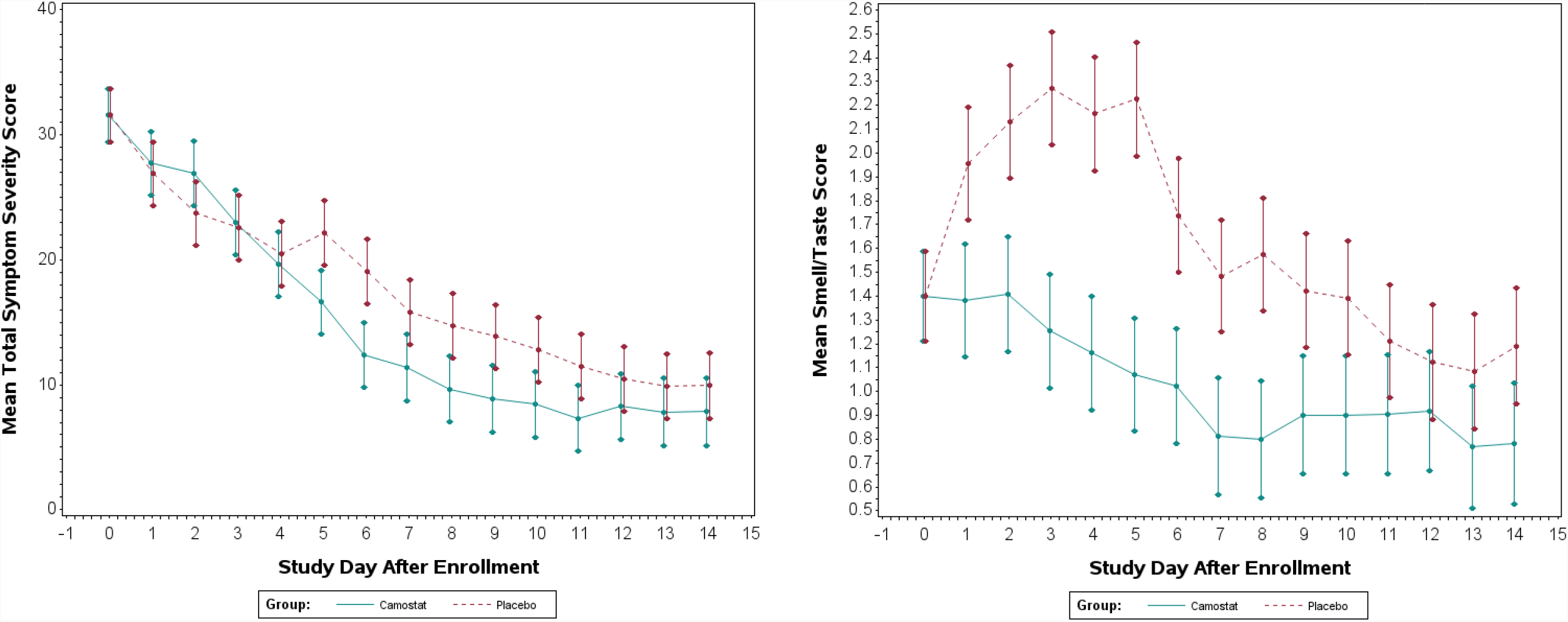
Comparison of patient-reported symptom scores in camostat vs. placebo groups

The complete symptom score data are provided in Supplementary Appendix.

### ADVERSE EVENTS

No study participant experienced progression to severe COVID-19 disease or death. Two patients experienced severe adverse events and were hospitalized during the study for worsening COVID-19 symptoms; one patient treated with placebo and one patient treated with camostat mesylate. Camostat was well tolerated and was associated with minimal side effects. Eleven (31.4%) patients treated with camostat experienced at least one adverse event compared to 6 (17.1%) patients treated with placebo.

## DISCUSSION

In this randomized, double-blind, placebo-controlled Phase 2 trial, a 7-day course of oral camostat mesylate given early in COVID-19 did not achieve the primary endpoint of reducing upper respiratory SARS-CoV-2 viral load compared to placebo. However, despite a relatively small sample size, we discovered a clinically-relevant effect of camostat on certain prespecified secondary outcomes. Using a high-resolution scoring system, FLU-PRO-Plus, that included quantification of the perception of smell and taste in addition to standard clinical symptomatology measures of influenza, we found that camostat mesylate accelerated overall COVID-19 symptom resolution and prevented the loss of smell and taste.

Alterations in smell and taste are common in COVID -19, and many affected people consider these symptoms to be debilitating.^10^ Such symptoms may take weeks or months to recover, if ever.^11^ This study showed that the development of these symptoms were significantly prevented by camostat, and represents a potentially important therapeutic benefit from camostat treatment.

Our results mirror—indeed exceed--the beneficial effects of oseltamivir in influenza^12-14^: while influenza viral load was not affected in the oseltamivir clinical trials, reduction of overall symptom scores was accelerated by the administration of drug (camostat, by several days, compared to ∼1 day by oseltamivir^12-14^). We adapted the detailed symptom score patient-reported observation instrument originally developed for influenza, FLU-PRO, to this camostat trial by incorporating one additional question regarding loss/alternation of smell and taste. This new instrument, FLU-PRO-Plus, enabled us to determine with fine clinical detail the effects of camostat in early SARS-CoV-2 infection while other trials did not find positive effects of camostat, either in late disease (or as informee via press release). While certain symptom domains in FLU-PRO-Plus such as eye, throat, chest and GI symptoms did not improve with camostat compared to placebo, the overall severity score and domain of smell/taste alteration did. Limitations of this trial include the relative small sample size which make it difficult to drawn firm conclusions from any subgroup analysis such as age, gender, co-morbidities, etc.

The results of this trial raises several key unanswered regarding camostat’s mechanism of action in improving the clinical course of COVID-19. First, while there was not a significant difference in viral load as determined by quantitative RT-PCR between camostat and placebo groups, we did not assess whether the viral load found represented infectious/viable virus. Camostat is predicted to inhibit viral entry^2,4^ so that it possible that the lack of difference in viral load in the two groups might be, at least in part, related to that mechanism. Because viremia is not a feature of COVID-19 and sampling sites other than the upper respiratoryy tract is not feasible in the outpatient clinical trial settting, it is difficult to assess systemic infection by viral load quantification. Second, camostat is a host-directed treatment aimed at inhibiting the cell-surface associated serine protease, TMPRSS2. It is likely that camostat has effects on other serine proteases, notably those found in the coagulation cascade as previously described ^15,16^.

Given that the measurement of d-dimers is an important biomarker in COVID-19, it is possible that the salutary clinical effect of camostat may be on the host response at the level of endothelial cell-coagulation cascade interactions. Further work is needed to explore mechanisms by which camostat may have a beneficial effect in various aspects of COVID-19.

The important patient-focused, patient-reported symptomatic outcomes of camostat mesylate in early COVID-19 point to a need for further clinical trial testing. Important outcomes include determination whether this drug reduces risk for severe disease and hospitalization; synergizes with direct antiviral drugs; might be useful in post-exposure prophylaxis; and might be applicability to modulating symptoms in long COVID-19. The potential importance of an oral treatment for early COVID-19, with an inexpensive, repurposed drug with few side effects, and which reduces the risk of complications cannot be overstated in the context of the ongoing pandemic.

## Data Availability

All data produced in the present study are available upon reasonable request to the authors

## ACKNOWLEDGMENTS

This investigator-initiated study was supported by Kenneth C. Griffin, the Prostate Cancer Foundation, the COVID-19 Early Treatment Fund, and the Harrington Discovery Institute, institutional funds from the Department of Internal Medicine at the Yale School of Medicine, and the Yale Center for Clinical Investigation. This study was partially supported by the United State Public Health Service grant, 5UL1RR024139. Ono Pharmaceuticals provided the study drug, FOIPAN, at no cost to the study but had no input into study design or the writing or content of the manuscript

We thank the participants in this study, and Dr. Howard Soule, Michael Milken, Steve Kirsch, Dr. Diana Wetmore, Dr. Donald Stanski and many others for their advice and insight during the course of this study. We are grateful to Dr. Brian Smith, Tesheia Johnson, Rhoda Arzoomanian, Amy Hummel and other staff members of the Yale Center for Clinical Investigation for enabling this study. We thank Dr. Greg Deye of NIAID and Dr. John Powers of George Washington University and NIH for discussions regarding FLU-PRO and FLU-PRO-Plus, and we are grateful for a license from Leidos Biomedical Resaerch, Inc. to use FLU-PRO-Plus in this work.

## FIGURES/TABLES

**Table 1. Baseline Patient Characteristics**

**<u>See accompanying Excel file</u>**.

*The complete dataset of baseline patient characteristics is available in Supplementary Appendix.

**Table 2. Summary of Adverse Events by Severity by Treatment Group**

**<u>See accompanying Excel file</u>**.

