## Supplementary data 2 for "A Phase 2 Randomized, Double-Blind, Placebo-controlled Trial of Oral Camostat Mesylate for Early Treatment of COVID-19 Outpatients Showed Shorter Illness Course and Attenuation of Loss of Smell and Taste"

**The Effect of Camostat Mesylate on COVID-19 Infection in Ambulatory Patients: An Investigator-Initiated Randomized, Placebo-Controlled, Phase IIa Trial**

**Protocol Number: 2000027971**

**National Clinical Trial (NCT) Identified Number: NCT04353284**

**STATISTICAL ANALYSIS PLAN**

**April 5, 2021**

**Version 1.0**

**LIST OF ABBREVIATIONS AND DEFINITIONS OF TERMS**

*Abbreviation	Definition*

AE	Adverse Event

ANOVA	Analysis of Variance

CEC	Clinical Events Committee

CI	Confidence Interval

DSMB	Data Safety Monitoring Board

eDC	Electronic Data Capture

ERC	Endpoints Review Committee

FDA	Food and Drug Administration

GEE	Generalized Estimating Equations

GCP	Good Clinical Practice

ICH	International Conference on Harmonization

IRB	Institutional Review Board

KM	Kaplan-Meier

MAR	Missing At Random

MCAR	Missing Completely At Random

MNAR	Missing Not At Random

PI	Principal Investigator

QOL	Quality of Life

R&D	Research and Development

SAE	Serious Adverse Event

SI	Site Investigator

YCAS	Yale Center for Analytical Sciences

1. ****PROTOCOL SYNOPSIS****

| **Objectives:** | Primary Objective: To determine whether camostat mesylate reduces SARS-COV-2 viral load in early COVID-19 disease.  Secondary Objective: To determine the effect of camostat mesylate on COVID-19 symptom score. |
| --- | --- |
| **Endpoints:** | Primary Endpoint:   1. Change (reduction) from baseline to day 4 in Nasopharyngeal swab and saliva viral load (RT-PCR log10).   Secondary Endpoints:   1. Change (reduction) from baseline to day 2 and to day 6 in Nasopharyngeal swab and saliva viral load (RT-PCR log10). 2. Difference in rate of a positive COVID-19 test result at day 6, 14 and 28 after enrollment 3. Change of COVID-19 symptom score from baseline to day 6 and to day 14 4. To assess the safety of camostat mesylate 200 mg po QID   Exploratory Endpoints:   1. Level of SARS-CoV-2 antigen from Nasopharyngeal specimens from baseline to days 6 and 14; there is no specific *a priori* power analysis for this exploratory analysis, however, the endpoint will provide mechanistic data regarding viral antigen levels after treatment with camostat mesylate. 2. To gain insight into whether clinical outcomes might improve after treatment with camostat mesylate 200 mg po QID but this study is not powered to assess progression to hospitalization or other severe disease. |
| **Population:** | Male and female outpatients 18 and older with RT-PCR-confirmed COVID-19 recruited and enrolled from study site or referred/presenting for the study because of a positive COVID-19 test from an outside clinical laboratory. |
| **Phase:** | 2 |
| **Description of Sites/Facilities Enrolling Participants:** | Academic Medical Centers |
| **Description of Study Intervention:** | Camostat mesylate 200mg po QID or placebo for 7 days |
| **Study Duration:** | 1 year |
| **Participant Duration:** | 4 weeks |

2. INTRODUCTION

This document describes the planned analyses of efficacy and safety data for Protocol Number: 2000027971, The Effect of Camostat Mesylate on COVID-19 Infection in Ambulatory Patients: An Investigator-Initiated Randomized, Placebo-Controlled, Phase IIa Trial. The data collected in this study will be used to evaluate the antiviral efficacy of Camostat Mesylate in subjects with a positive COVID-19 test as determined by quantitative RT-PCR (qRT-PCR)***.*** Results from this endeavor will assist in establishing an efficacious approach for the treatment of COVID-19 positive patients***.***

A formal protocol and amendments were submitted for approval to the Central and the local Institutional Review Boards (IRB) at participating centers. The data analysis described follows the final protocol dated **August 18, 2020 Version 8.0**.

The outlined analytic plan is prospective in nature and is based upon the protocol. Protocol changes and amendments that might be introduced during the operation of the study will be included and appended to this analysis plan in chronological order. The effect of such changes/amendments on data analysis will be clearly described and assessed.

A single randomization scheme for 200002797 was generated by the Yale Center for Analytical Sciences (YCAS). This scheme was written in SAS version 9.4 (Cary, NC) with a 1:1 allocation of Camostat to placebo, using a permuted block design. The program used to generate the randomization scheme is available from the YCAS. Each participant in the study will have a documented study entry/enrollment date. Enrollment reports will be available on a regular basis, both by site and overall.

The analysis will include all study participants who have contributed data to the study. The study report will include a full accounting of all persons randomized and enrolled in the study.

Paper worksheets are used for collection of baseline characteristics, follow-up data and other variables that will be entered in electronic data capture forms (eDC) and used in the analysis phases of the study; additional data for clinical outcomes will be extracted from the electronic health record (EHR). eDC forms have been developed, tested and validated prior to their use. All enrolling sites have access to the trial management system for enrolling participants and entering data.

This study will also assess safety: adverse events and death information will be collected. For patients who drop out or are lost to follow-up, a date and reason, if available, will be collected on the termination/study exit data form.

3. STUDY OBJECTIVES AND ENDPOINTS

| OBJECTIVES | ENDPOINTS | JUSTIFICATION FOR ENDPOINTS |
| --- | --- | --- |
| Primary |  |  |
| 1. To determine whether camostat mesylate reduces SARS-COV-2 viral load within 2-3 days after laboratory-confirmed diagnosis of COVID-19 disease in ambulatory patients. | 1. Primary Endpoint: Change (reduction) from baseline to day 4 in respiratory (Nasopharyngeal swab, saliva RT-PCR) log_10_ viral load . | Per FDA recommendations and our own judgment, the timing of this primary endpoint was chosen because  reduction of viral load in respiratory tract samples would be expected to be associated with anti-viral activity of the study drug, and would be expected to be associated with continued reduction of viral load during drug administration, hence potential reduction severity of COVID-19 disease and reduction of transmission by reducing infectiousness of infected individual.  If we are able to achieve a reduction in viral load then a trial with clinical endpoints such as risk for progression to hospitalization and/or need for intensive unit care would be justified. |
| Secondary |  |  |
| 1. To determine whether camostat mesylate reduces SARS-COV-2 viral load within 7 days after laboratory-confirmed diagnosis of COVID-19 disease in ambulatory patients.  2. To determine the effect of camostat mesylate on COVID-19 symptom score.  3. To evaluate the safety of camostat mesylate  4. Adverse events | 1. Change from baseline to day 2 and to day 6 in respiratory (Nasopharyngeal swab, saliva RT-PCR) log_10_ viral load.  2. Difference in rate of positive COVID-19 test at day 6, 14 and 28  3. Change of COVID-19 symptom score from baseline, to days 6 and 14  4. Assessment of safety by labs and history and physical, according to The Common Terminology Criteria for Adverse Events  (CTCAE) Version 5.0. | As for the primary endpoints (above).  We expect to see a time-dependent reduction of viral load, hence the second time point.  We expect to see a reduction in symptom score from baseline at day 6 and day 14 +/-2.  If the secondary endpoints are not achieved, then a trial to examine the effect of camostat mesylate on COVID-19 would not be justified. |
| Tertiary/Exploratory |  |  |
| - 1. To determine clinical outcome of COVID-19 treated with camostat mesylate - 2. To determine conversion of SARS-CoV-2 antigen from positive at baseline to negative at days 6 and 14. | 1. As an exploratory endpoint in comparing treatment to placebo groups, we will assess time to clinical improvement and risk for hospitalization*.*  2. Quidel’s Sofia assay will be used for antigen detection. | A tertiary objective not powered by this study. We will explore, however, whether the treatment group clinically improves or has a lower, neutral, or greater risk of hospitalization.  As the Quidel assay has not been used for serial assessment of outpatients, this exploratory analysis will provide data regarding the performance of this test in our outpatient context |

4. PROTOCOL AMENDMENTS

During the course of the trial the following protocol amendments have been made:

| **Version** | **Date** | **Description of Change** | **Brief Rationale** |
| --- | --- | --- | --- |
| 6.0 | May 21, 2020 | N/A – first version approved by IRB | N/A |
| 7.1 | June 11, 2020 | Increase dosing from TID to QID and change in primary endpoint evaluation from day 2 to day 4  Operational changes as a result of institutional changes at Yale (eg different clinic)  Clarifications of procedures | Feedback from FDA  Yale institutional operational refinements |

5. INVESTIGATORS AND SITES

A total of **1** site is participating in the trial to accrue ***114*** patients. Thus, each site is expected to enroll a total of **114** patients. Patient intake for the trial is scheduled to last for ***1*** year, with each site expected to enroll an average of ***114*** patients per year.

The study is expected to last a total of **1** year. Table **5.1** provides a list of the participating sites and site investigators.

**Table 5.1. List of participating sites and site investigators.**

| **Site Location** | **Site Investigator** |
| --- | --- |
| Yale | Geoffrey Chupp, MD |

6. INVESTIGATIONAL PLAN
   6.1 Study Design

This is a randomized, double-blind, placebo-controlled phase II trial comparing camostat mesylate to placebo in adults with SARS-CoV-2 infection as determined by a validated test who are receiving standard supportive care in an outpatient setting.

The 2 arms to be compared in the trial are:

- Camostat mesylate 200 mg PO QID for 7 days
- Placebo 200 mg PO QID for 7 days

This study will be conducted in approximately 114 adult subjects ages ≥ 18 who are SARS-CoV-2 positive as determined by a validated test and within 3 days of being notified of their first positive COVID-19 test result. The study will enroll subjects who have at least one of the following COVID19-related symptoms indicating mild disease: fever upper respiratory symptoms, cough, chills, loss of taste/smell or a recent high-risk exposure to COVID-19. Subjects will be treated with 200 mg camostat mesylate PO, QID, or with placebo PO, QID for 7 days. The treatment schedule is outlined in the Schedule of Activities (SoA), Table 1. Subjects will be followed at 2, 4, 6, 11, 14 and 28 days after the treatment start.

Participants will be randomized equally to camostat mesylate or identical appearing placebo using a permuted-block design with variable block size. Actual treatment assignment will be concealed from the investigators and the participants.

The primary hypothesis to be tested is that SARS-CoV-2 clearance as determined by quantitative RT-PCR (qRT-PCR) from nasopharyngeal samples at day 4 is greater (i.e. greater reduction in viral load) in camostat mesylate compared to placebo.

1 enrolling sites (Yale) is expected to participate in order to accrue ***112*** patients.
  6.2 Study Schema

Total N=114: Screen potential participants by inclusion and exclusion criteria; obtain relevant history, document all contacts with potential participants; refer to study site.

Prior to

Enrollment

Visit 1

**At designated facility: Informed consent, randomization (after urine pregnancy test as indicated)**

**A. Perform baseline assessments, study symptom assessment plus patient COVID-19-PRO daily for days 0-14, 28)**

**B. Specimens to be collected**. **Nasopharyngeal swabs (2), saliva; research labs blood for CBC, chemistry, urine pregnancy test, research labs including serum, PBMC isolation, coagulation component measurements**

**C. Provide 2 d of Camostat** **(8 x 200 mg capsules or 8 placebo capsules) for days 0 and 1; 200 mg po qid or matched placebo**

Day 0

At in-person Visit 1 on Day 0: Randomize 1:1 treatment (arm A) vs. placebo (arm B)

Visits 2-4

**At designated facility**

**A. Perform followup assessments, days 2, 4, 6 in person (study symptom assessment plus patient COVID-19-PRO)**

**B. Specimens to be collected**. **On day 4: required** **Nasopharyngeal swabs (2), saliva; on days 2 and 6, NP swabs optional but saliva required**

**C. Provide 2 d of Camostat (8 x 200 mg capsules or 8 placebo capsules) for days 2, 3 and 4, 5 and 6, 7; 200 mg po qid or matched placebo**

Days 2, 4, 6

Visit 5

**At designated facility**

**A. Perform followup assessment in person (study symptom assessment plus patient COVID-19-PRO)**

**B. Specimens to be collected**. **Nasopharyngeal swabs (2), saliva; research labs blood for CBC, chemistry, urine pregnancy test, research labs including serum, PBMC isolation, coagulation component measurements**

Day 14+/-2

Visit 6

**FINAL ASSESSMENT**

**At designated facility**

**A. Perform followup assessment, in person (study symptom assessment plus patient COVID-19-PRO)**

**B. Specimens to be collected**. **Nasopharyngeal swabs (2), saliva; research labs blood for CBC, chemistry, urine pregnancy test, research labs including serum, PBMC isolation, coagulation component measurements**

Day 28+/-2

6.3 Study Cohort

This clinical trial can fulfill its objectives only if appropriate participants are enrolled. The protocol-specified eligibility criteria are designed to select subjects for whom study participation is considered appropriate. All relevant medical and non-medical conditions should be taken into consideration when deciding whether this protocol is suitable for a study candidate. Eligibility criteria may not be waived by the investigator and conformance to the eligibility criteria will be reviewed in the case of a GCP or a regulatory authority audit. Any questions regarding a study candidate’s eligibility should be discussed with the medical monitor before enrollment. Reference is made to the National Early Warning Score (NEWS) for grading of illness intensity and the Common Terminology Criteria for Adverse Events (CTCAE), Version 5.0 for severity of adverse events.

6.3.1 Inclusion Criteria

Consecutive participants testing positive for COVID-19 at the Sargent Drive testing center, any other YNHHS site, or external COVID-19 testing facility will be eligible for enrollment.

In order to be eligible to participate in this study, an individual must meet all of the following criteria:

1. Be enrolled within 3 days of being notified of their first positive COVID-19 test result.
2. Evidence of a recent active COVID-19 infection, as evidenced by the positive test results being associated with at least one COVID-19-compatible symptom such as fever, upper respiratory symptoms, cough, chills, loss of taste/smell, etc.(see COVID-19-PRO symptom score sheet), or a recent high-risk exposure to COVID-19
3. Provision of informed consent
4. Stated willingness to comply with all study procedures and availability for the duration of the study
5. Male or female, aged 18 and older
6. Diagnosed with COVID-19 within past 3 days and not exhibiting manifestations requiring hospitalization such as extreme shortness of breath or severe prostration. Nurses at the study site will assess such severe conditions requiring hospitalization, which would preclude enrollment.
7. Ability to take oral medication and be willing to adhere to the camostat mesylate regimen.
8. For men and women of reproductive potential: use of condoms or other methods to ensure effective contraception with partner during study drug administration.
9. Agreement to adhere to Lifestyle Considerations (birth control measures) throughout the 7-day duration of study drug administration.
10. English and Spanish speaking subjects as well as patients speaking any language for which we can find appropriate translators will be enrolled. A short form with interpretation will be used for anyone speaking a language for which a translated informed consent form is not currently available in accordance with local site IRB policies, including developing certified translations as necessary.
    6.3.2 Exclusion Criteria

Study candidates who meet any of the following criteria will not be eligible for participation in this study:

An individual who meets any of the following criteria will be excluded from participation in this study:

1. Presence of COVID-19 disease manifestations that would require referral for consideration of hospitalization.
2. A previous positive COVID-19 test reported more than 7 days before, which would indicate likelihood of non-culturable, nonreplicating virus
3. A positive COVID-19 test without a known recent exposure that would indicate an active infection, hence an unknown chance of non-culturable, non-replicating virus being present (i.e., asymptomatic COVID-19 infection of unknown duration)
4. Known pregnancy or lactation; a positive urine pregnancy test done at the enrollment (day 0 visit) for women with child bearing potential.
5. Known allergic reactions to components of camostat mesylate.

With regard to inclusion or exclusion of women of child-bearing potential, women who tell us they know they are pregnant are excluded. All women of child-bearing potential who test positive for pregnancy by urine test at first visit are excluded. A day 14 followup blood pregnancy test will be done on appropriate enrolled women (i.e. those who had a negative urine pregnancy test on day 0 for further safety assessment.

6.4 Blinded Study Drug (camostat mesylate and placebo)
    6.4.1 Description

Camostat mesylate will be provided by Ono Pharmaceuticals as 500 tablets/bottle in glass bottles. Each tablet contains 100 mg of Camostat mesylate. For study administration two tablets will be combined (by investigational pharmacy) into one 200 mg capsule and subjects will take one capsule qid of either study drug or placebo. Placebo will be formulated to be similar in appearance to the study drug and provided in similar plastic containers.

Chemical name: 4-[[4-[(Aminoiminomethyl)amino]benzoyl]oxy]benzeneacetic acid 2- (dimethylamino)-2-oxoethyl ester methanesulfonate.

International Nonproprietary Name (INN): Camostat

Molecular formula: C_20_H_22_N_4_O_5_ · CH_4_O_3_S

Molecular weight: 494.52

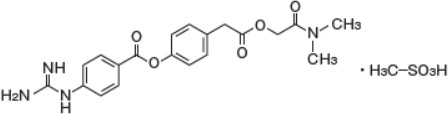
Structural formula

6.4.2 Source

Ono Pharmaceutical, Japan, will provide Camostat mesylate 100 mg tablets as FOIPAN. The Yale New Haven Hospital research pharmacy will receive and store the drug within 15-30°C range according to protocol storage requirements.

Microcrystalline Cellulose NF (PH-102) for placebo formulation will be acquired from Fagron.

Empty gelatin capsules Size 0, for over-encapsulation will be acquired from Fagron.

Before Ono Pharmaceutical can send the drug, an IND will be obtained from the FDA.

6.4.3 ****Packaging and Labelling****

Encapsulated Camostat 200 mg Capsules

For blinding, two intact active camostat 100 mg tablets will be placed in a capsule shell, back filled with sufficient quantity of microcrystalline cellulose, and closed. The capsules are visually inspected and packed into polypropylene bottles.

Placebo for Encapsulated Camostat 200 mg dose Capsules

Matching placebo will be compounded, using a matching empty capsule filled with sufficient quantity of microcrystalline cellulose. The capsules are visually inspected and packed into polypropylene bottles.

Assigned beyond use date is not later than the time remaining until the earliest expiration date of any ingredient or 6 months.

Master formulation record and compounding record will be kept for the compounding. Compounding, packaging and labelling will be carried out by Investigational Drug Service Pharmacy, Yale-New Haven Hospital.

*Labeling:*

Camostat 200 mg dose Capsules (Active)	Compounding Date:

Lot:	Exp Date:

Qty:

Protocol #

Storage Condition (Room Temp 15-30°C)

"Caution: New Drug--Limited by Federal (or United States) law to investigational use."

Placebo for Camostat 200 mg Capsules	Compounding Date:

Lot:	Exp Date:

Qty:

Protocol #

Storage Condition (Room Temp 15-30 °C)

"Caution: New Drug--Limited by Federal (or United States) law to investigational use."

6.4.4 ****Shipping, Storage, and Stability****

The study drug Camostat is stable at 25°C and should be kept at ambient (15-30°C) temperature for long term storage. For short term storage (up to 30 days) to facilitate daily dispensing of the drug it will be kept at ambient office or clinic temperature.

Placebo for Camostat will also be stored at ambient (15-30°C) temperature for long term storage. For short term storage (up to 30 days) to facilitate daily dispensing of the drug it will be kept at ambient office or clinic temperature.

Drug will be stored in locked cabinets at ambient (15-30°C) temperature at all times other than during active dispensing.

6.4.5 ****Dispensing****

A pharmacist or other qualified staff member will dispense bottles containing study drug. For dispensing, 8 capsules of overencapsulated Camostat 200 mg per capsule or identically appearing Placebo for camostat mesylate 200 mg will be packaged in a polypropylene bottle and given to enrolled research participants every other day. The bottles will be labeled with dispensing label containing patient name, administration instructions, and provider information per state pharmacy regulations.

6.4.6 ****Return and Compliance Assessment****

After the completion of the treatment course, empty, partially used, or full bottles of study drug will be retrieved from the subject. The quantities of unused study drug and the date when these study supplies are returned by the subject should be recorded in the study drug accountability records. In addition, the subject’s dosing diary should be reviewed, any incomplete or inconsistent entries should be addressed, and the dosing diary should be copied. Returned capsules and bottles may be destroyed according to the site’s standard operating procedures.

6.4.7 ****Accountability****

The Sponsor-Investigator and the Yale New Haven Hospital research pharmacy will keep accountability records for all investigational products acquired, dispensed, used and disposed.

.

6.4.8 ****Emergency Unblinding****

Every attempt should be made to preserve the integrity of study drug blinding. Unblinding in individual subjects who experience adverse events is rarely required to provide effective intervention and support. For subjects having adverse events or laboratory abnormalities that require drug cessation or medical intervention, the investigational staff should strive to provide necessary support to the subject without breaking the blind. In the exceptional circumstances that knowledge of the study drug assignment appears essential for providing appropriate medical management, the investigator should contact the medical monitor to discuss the rationale for breaking the blind and the adverse consequences of the unblinding for the subject’s continued participation in the study. If the investigator still believes that unblinding is warranted, the investigator will be able to access the randomization system in order to obtain the treatment assignment for that subject. No randomization lists or other unblinding information will be provided to the site. After breaking the blind, the investigational site staff should record details regarding the reasons for breaking the blind and any adverse events leading to the breaking of the blind in the source documents and in the appropriate eCRF.

If the site breaks the blind, that subject will be discontinued from study therapy.

6.5 Randomization

Patients must consent for randomization. All pre-randomization evaluation will be completed prior to randomization.

After a subject has completed the necessary screening assessments and has been confirmed to be eligible, the subject can be randomized into the study. In order to obtain a treatment arm assignment for a subject, a site representative will submit the appropriate information for randomization, and the system, will make an assignment of a study drug bottle number.

Subjects will be randomized in a 1:1 ratio to either of the following treatment assignments:

- Camostat mesylate
- Placebo

In order to balance treatment allocation, a permuted block centralized randomization will allocate subjects. The randomization scheme will be computer-generated, and the allocation sequence will be concealed.

It is anticipated that subjects will usually begin taking study drug immediately after randomization Day 0 of the study. Once assignment is made, it is final and the patient will be analyzed in that group regardless of future events or information, in accordance with the intent-to-treat principle.

6.6 Blinding

The identity of the treatments will be concealed by central blinding of study drug assignments. Blinding will be accomplished through use of a placebo that is well-matched to the active drug in appearance, packaging, labeling, and schedule of administration. During the study, subjects, caregivers, investigational site personnel, study sponsor study team members, and all other study personnel will remain blinded to the identity of the treatment assignments; these assignments will be available only to the unblinded randomization team, the DSMB for the study (if requested), and drug safety personnel who are not part of the study team. Unblinding during the study will only occur in the case of subject emergencies or as requested by the DSMB. Where required by local regulation, expedited reporting of serious adverse events to specific regulatory authorities will include information regarding the study drug treatment assignment; this will be done in such a way that subjects, investigational site personnel, institutional review board/independent ethics committees (IRB/IEC), and study team members remain blinded as to the treatment assignment for the subject described in the adverse event report.

The final unblinded analysis of the study will only be performed when the database is completed and locked. Except for emergency unblinding, individual subjects, caregivers, and site personnel will not be informed of the randomized treatment assignments until the implications of revealing such data for the overall camostat study program have been determined by the project leader for the camostat development program.

6.7 Reporting of Adverse Medical Events

An adverse event (AE) is defined as any adverse change from the patient's baseline conditions, including any significant clinical or laboratory test value abnormality that occurs during the course of the study, regardless of its association with the study procedures.

For purposes of this trial, the AE data collection mechanism is as follows:

AEs will be described using concise medical terminology, including whether the AE is serious, the date of AE onset, the date of AE resolution, the severity of the AE based on the CTCAE, Version 5.0, a description of relatedness of the AE to study drug or to a study procedure, the action taken due to the AE, and the outcome of the AE. AEs will be assessed during Screening and on each designated day. Day 22 assessments will be done via telephone. AEs will be coded in and reported by MeDRA (Medical Dictionary for Regulatory Affairs) version 23.0.

An AE is any untoward medical occurrence in a clinical investigation subject administered a medicinal product; the event does not necessarily have a causal relationship with study drug administration or usage. An AE can therefore be any unfavorable and unintended sign (including an abnormal laboratory finding), symptom, or disease temporally associated with the use of a medicinal product, whether or not considered related to the medicinal product.

In this study, any of the following events will be considered an AE:

- Any complication that occurs as a result of a protocol-mandated procedure (e.g. venipuncture, ECG).
- Any preexisting condition that increases in severity or changes in nature during or as a consequence of study drug administration. Worsening manifestations of the underlying infection (e.g. increase in COVID-19 manifestations) may be considered AEs in this study.
- Any injury or accident. If a medical condition is known to have caused the injury or accident (e.g. a fall secondary to dizziness), the medical condition (dizziness) and the accident (fall) should be reported as 2 separate AEs.
- Any abnormality in physiological testing or a physical examination finding that requires clinical intervention or further investigation (beyond ordering a repeat [confirmatory] test).
- Any laboratory (e.g. clinical chemistry, hematology, urinalysis) or investigational abnormality (e.g. ECG, X-ray) independent of the underlying medical condition that requires clinical intervention, results in further investigation (beyond ordering a repeat [confirmatory] test), or leads to investigational medicinal product interruption or discontinuation unless it is associated with an already reported clinical event. If the laboratory abnormality is part of a syndrome, the syndrome or diagnosis (e.g. anemia) not the laboratory result (e.g. decreased hemoglobin) should be recorded.
- A complication related to pregnancy or termination of a pregnancy (see Protocol for additional information).

None of the following events is considered an AE:

- Laboratory abnormalities not requiring clinical intervention or further investigation. Such abnormalities will be captured as part of laboratory monitoring.
- A diagnostic, medical, or surgical procedure (e.g. surgery, endoscopy, tooth extraction, transfusion). However, the medical condition for which the procedure was performed should be reported if it meets the definition of an AE. For example, an acute appendicitis that begins during the AE reporting period should be reported as the AE and the resulting appendectomy should be recorded in the source documents.
- A preexisting disease or condition or laboratory abnormality present or detected before the initial screening visit and that does not worsen.
- An intervention not associated with an untoward medical occurrence (e.g. hospitalization for elective surgery or for social and/or convenience reasons).
- An overdose without clinical sequelae.
  6.7.1 Serious Adverse Events

An SAE is defined as an untoward medical occurrence that results in any of the following outcomes:

- Death (i.e., all deaths occurring between signing of the consent form to within 30 days after last study drug administration), including deaths due to COVID-19 if no other event more satisfactorily explains the reason for death. Deaths that occur as a result of an AE that started during the study period should be reported. Death is not an SAE term; the reported AE should be the event that caused the death. Death is the outcome of this SAE.
- Life-threatening situation (ie, with an immediate risk of death from the event as it occurred but not including an event that, had it occurred in a more serious form, might have caused death).
- In-patient hospitalization or prolongation of existing hospitalization. Of note, an untoward medical occurrence that occurs during hospitalization is an AE but a complication that prolongs hospitalization is an SAE. In-subject hospitalization comprises formal admission to a hospital for medical reasons, for any length of time, whether or not hospitalization extends overnight. However, hospital admissions or prolongations of hospitalization for administration of the study drug or procedures required by the study protocol, diagnostic observations or procedures, administration of concomitant or subsequent therapy for the cancer, logistical issues (e.g. lengthy travel), or the convenience of the subject or clinical personnel are not considered serious.
- Persistent or significant disability/incapacity.
- Congenital anomaly/birth defect in the offspring of a subject who received the investigational medicinal product.
- Other medically significant event. Such events may not be immediately life-threatening or result in death or hospitalization, but based upon appropriate medical and scientific judgment, may jeopardize the subject, or may require medical or surgical intervention to prevent one of the outcomes listed above. Examples of such events might include:
  - Allergic bronchospasm requiring intensive treatment in an emergency room or at home
  - New cancers or blood dyscrasias
  - Convulsions that do not result in hospitalization
  - Development of drug dependency or drug abuse

For this trial, adverse events meeting the definition of serious will be collected on all patients, regardless of their association with the study treatment, from the time of randomization to the end of a patient’s follow-up.

6.7.2 Unexpected Adverse Event

An unexpected AE is defined as an event that has a nature, severity, or specificity that is not consistent with the applicable investigator brochure, or that is symptomatically and pathophysiologically related to a known toxicity but differs because of greater severity or specificity. For example, under this definition, hepatic necrosis would be unexpected (by virtue of greater severity) if the investigator brochure only referred to elevated hepatic enzymes or hepatitis. Similarly, cerebral thromboembolism and cerebral vasculitis would be unexpected (by virtue of greater specificity) if the investigator brochure only listed cerebral vascular accidents. “Unexpected,” as used in this definition, refers to an adverse drug experience that has not been previously observed and reported rather than an experience that has not been anticipated based on the pharmacological properties of the study drug.

6.7.3 Treatment-Emergent Adverse Event

A Treatment-Emergent Adverse Event (TEAE) is defined as an AE that occurs or worsens in the period from the first dose of study drug administration to Day 28 of the study.

6.7.4 Adverse Events of Special Interest

There are no adverse events of special interest in this protocol.

6.7.5 Eliciting Adverse Event Information

The investigator is to report all directly observed AEs and all AEs spontaneously reported by the study subject. In addition, each study subject should be questioned about AEs at each scheduled study center visit or during any telephone contact with the subject. The type of question asked should typically be open-ended, e.g. “Have you had any new health problems?” or a similar type of query.

6.7.6 Recording Adverse Events

All AEs will be assessed by the investigator or qualified designee and recorded in the electronic case report forms (eCRFs). The following considerations apply in collecting AE information:

- Recognized medical terms (not colloquialisms, abbreviations, or jargon) should be used when recording AEs. Ideally the description should be consistent with MedDRA nomenclature.
- The medical diagnosis (i.e., disease or syndrome) should be recorded instead of signs and symptoms (e.g. pneumonia instead of shortness of breath, coughing, and fever).
- Any sign or symptom considered as unrelated to an encountered disease or syndrome should be recorded as an individual AE (e.g. if nausea and severe headache are observed at the same time, each event should be recorded as a separate AE).
- If an AE resolves and reoccurs at a later date, a new AE must be documented and reported, as appropriate.

The following information should be recorded:

- Description as to whether the AE is serious, if applicable (see Protocol)
- The start date (date of AE onset)
- The stop date (date of AE resolution)
- The severity of the AE (see Protocol)
- A description of the relatedness of the AE to the study drug or to a study procedure (see Protocol)
- The action taken due to the AE
- The outcome of the AE
  6.7.7 Grading of the Severity of an Adverse Event

The severity of AEs will be graded and reported using the CTCAE, Version 5.0.

If a CTCAE criterion does not exist, the investigator should use the grade or adjectives: Grade 1 (mild), Grade 2 (moderate), Grade 3 (severe), Grade 4 (life-threatening), or Grade 5 (fatal) to describe the maximum intensity of the AE. For purposes of consistency with the CTCAE, these intensity grades are defined in Table 6.7.1.

Table 6.7.1.	Grading of Adverse Event Severity

| **Grade** | **Adjective** | **Description** |
| --- | --- | --- |
| Grade 1 | Mild | Sign or symptom is present, but it is easily tolerated, is not expected to have a clinically significant effect on the subject’s overall health and well-being, does not interfere with the subject’s usual function, and is not likely to require medical attention. |
| Grade 2 | Moderate | Sign or symptom causes interference with usual activity or affects clinical status and may require medical intervention. |
| Grade 3 | Severe | Sign or symptom is incapacitating or significantly affects clinical status and likely requires medical intervention and/or close follow-up. |
| Grade 4 | Life-threatening | Sign or symptom results in a potential threat to life. |
| Grade 5 | Fatal | Sign or symptom results in death. |

The distinction between the seriousness and the severity of an AE should be noted. Severity is a measure of intensity; thus, a severe reaction is not necessarily a serious reaction. For example, a headache may be severe in intensity, but would not be classified as serious unless it met one of the criteria for serious events (as listed in Protocol).

6.7.8 Describing Adverse Event Relationship to Study Drug and Study Procedures

The investigator will evaluate the causal relationship of each AE to a study drug and/or to a study procedure (e.g. venipuncture or ECG evaluation) and record that relationship on the appropriate CRFs. Causality will be assessed considering whether the AE is reasonably related to the study drug or procedure or whether the AE is not reasonably related to the study drug or procedure considering the definitions in Table 6.7.2.

It should be emphasized that the determination of the relationship of an adverse event to study drug should be based on the presumption that the study subject is receiving active drug even though the blind remains unbroken. As noted in Protocol, unblinding should be avoided unless absolutely necessary for a subject’s well-being; breaking the blind should not be performed merely to obtain enhanced certainty regarding the relationship of an adverse event to study drug.

| **Table 6.7.2 Relationship of Study Drug to Adverse Event** | |
| --- | --- |
| **Relationship** | **Description** |
| Definite | A clinical event in which a relationship to the use of the study drug seems definite because of such factors as consistency with known effects of the drug; a clear temporal association with the use of the drug; lack of alternative explanations for the event; improvement upon withdrawal of the drug (de-challenge); and recurrence upon resumption of the drug (rechallenge). |
| Probable | A clinical event in which a relationship to the study drug seems probable because of such factors as consistency with known effects of the drug; a reasonable temporal association with the use of the drug; lack of alternative explanations for the event; and improvement upon withdrawal of the drug (de-challenge). |
| Possible | A clinical event with a reasonable temporal association with administration of the study drug, and that is not likely to be explained by concurrent disease or other drugs or chemicals. Information on drug withdrawal may be lacking. |
| Unlikely | A clinical event with a temporal relationship to study drug administration that makes a causal relationship improbable and for which other factors suggesting an alternative etiology exists. Such factors might include a known relationship of the clinical event to a concomitant drug, past medical history of a similar event, the subject’s disease state, intercurrent illness, or environmental factors. |
| Unrelated | A clinical event in which a relationship to the study drug seems improbable because of factors such as inconsistency with known effects of the study drug; lack of a temporal association with study drug administration; lack of association of the event with study drug withdrawal or rechallenge; and/or presence of alternative explanations for the event. Alternative explanations might include a known relationship of the clinical event to a concomitant drug, past medical history of a similar event, the subject’s disease state, intercurrent illness, or environmental factors. |

6.7.9 Adverse Event Reporting Period

The start of the AE reporting for a study subject will coincide with the first administration of study drug for this study. After the first administration of study drug, all AEs (serious and nonserious, related and unrelated) should be reported. Unless a subject withdraws consent for follow-up, each subject must be followed until the end of the AE reporting period at Day 28 or when any ongoing drug-related AEs and/or SAEs have resolved or become stable. The investigator should use appropriate judgment in ordering additional tests as necessary to monitor the resolution of events. The medical monitor or the sponsor may request that certain AEs be followed longer.

Investigators are not obligated to actively seek information regarding the occurrence of new SAEs beginning after Day 28. However, if the investigator learns of such an SAE and that event is deemed relevant to the use of LAM‑002A, he/she should promptly document and report the event. A longer reporting period applies in the case of pregnancy (see Protocol).

6.7.10 Study Center Reporting Requirements

### 6.7.10.1 Adverse Event Reporting Requirements

Classification of an event as serious or nonserious (see Protocol) determines the reporting procedures to be followed by the study center. Study center reporting requirements for AEs are summarized in Table 6.7.3.

| **Classification** | **Reporting Time** | **Reporting Action** |
| --- | --- | --- |
| Serious | Within 24 hours of becoming aware of the event | E-mail report on designated SAE report form to the sponsor or designee [a] and to the study center IRB, as per local IRB/IEC requirements |
|  | Within 10 working days | E-mail copies of relevant source documents (e.g. progress notes, laboratory and diagnostic test results, discharge summaries) [b] to the sponsor or designee [a]. |
|  | Per eCRF submission procedure | Record and submit information on appropriate eCRFs. |
| Nonserious | Per eCRF submission procedure | Record and submit information on appropriate eCRFs. |
| a. See contact information in Pharmacy Manual. | | |
| b. Subject name, address, and other personal identifiers should be obscured but without losing the traceability of a document to the study subject identifiers. | | |
| **Abbreviations:** eCRF=case report form; IRB/IEC= institutional review board/independent ethics committee; SAE=serious adverse event | | |

Table 6.7.3.	Study Center Reporting Requirements for Adverse Events

For SAEs, in addition to completing the AE portion of the eCRF, an SAE report form must also be completed and emailed within 24 hours of first awareness of the event to the sponsor (or designee).

Investigators must also submit written safety reports as required by the IRB/IEC within timelines set by local and regional regulations. The study site should retain documentation of the submission of expedited safety reports to the IRB/IEC, and their receipt. Study site personnel should not wait to receive complete information or the investigator’s signature before notifying the sponsor (or designee) and the IRB/IEC of an SAE.

The following minimum information is required:

- Subject identification (i.e. subject number, sex, age)
- Description of the SAE (diagnosis preferred, symptoms, etc.)
- Study drug and causal relationship of the SAE to the study drugs or study procedures
- Investigator name

Collection of complete information concerning SAEs is extremely important. The information in the AE portion of the eCRF and the SAE report form must match or be reconciled. Where the same data are collected, the forms must be completed in a consistent manner. For example, the same AE term should be used on both forms.

All available and relevant supporting documentation (e.g. progress notes, pertinent laboratory and diagnostic test results, discharge summaries) should also be provided. The subject’s name, address, and other personal identity information should be obscured on any source documents (e.g. progress notes, nurses’ notes, laboratory and diagnostic test results, discharge summaries). The subject’s unique study number should be included on each page of any document provided.

The sponsor (or designee) will review SAE reports for missing information and send queries to the site for resolution as indicated. The study site is responsible for obtaining pertinent follow‑up information missing from initial reports and forwarding the information within 24 hours of receipt to the sponsor (or designee). Follow‑up information to the SAE should be clearly documented as “follow-up” in the SAE report form.

The original SAE form and any follow‑up forms must be kept on file at the study site. An SAE is followed until it is considered resolved, it returns to baseline, is chronically ongoing, or explained otherwise by the principal investigator.

### 6.7.10.2 Contact Information for Reporting an SAE or Pregnancy

This will be managed by Yale Center for Clinical Investigation (YCCI).

6.7.11 Sponsor Reporting Requirements

Each SAE or special situation report received from the investigator will be evaluated by the sponsor (or designee). The sponsor (or designee) will assess the seriousness of the event, the expectedness of the event, and the relationship to participation in the study. The sponsor (or designee) will also indicate whether there is concurrence with the details of the report provided by the investigator.

The sponsor (or designee) will provide information for reporting of suspected, unexpected, serious adverse reactions (SUSARs) to regulatory authorities worldwide consistent with relevant legislation or regulations, including the applicable US FDA CFR, the European Commission Clinical Trials Directive (2001/20/EC, and revisions), and other country-specific legislation or regulations. SUSARs will be reported to regulatory authorities within 7 calendar days for life threatening and fatal events, or 15 calendar days for all others. These timeframes begin with the first notification of the event from the reporting investigator to the sponsor (or designee), which represents the start of the regulatory clock (Day 0).

The sponsor (or designee) will also provide all investigators with a safety letter describing the SUSAR. The information will be provided by e‑mail, fax, or overnight mail within 15 calendar days from Day 0. Investigators will be requested to provide written notification of the event to the relevant IRB/IEC as soon as is practical and consistent with local regulatory requirements and local institutional policy.

6.7.12 Study Intervention Discontinuation and Participant Discontinuation/Withdrawal
       6.7.12.1 **Discontinuation of Study Intervention**

Reasons why a study subjects may be withdrawn from the study intervention include, but are not limited to:

- Subject request (withdrawal of consent)
- Protocol violation, i.e. declining to take drug or be swabbed or providing saliva for viral load two times.
- AE or reactions
- Any condition, interaction, or contraindication where continued participation in the study will result in an unacceptable risk for the subject, as assessed by the Investigators or advisers
- Discontinuation of the study by the Sponsor
- Lost to follow-up
  6.7.12.2 **Participant Discontinuation/Withdrawal From the Study**

Participants are free to withdraw from participation in the study at any time upon request.

An investigator may discontinue or withdraw a participant from the study for the following reasons:

- Pregnancy
- Significant study intervention non-compliance
- If any clinical adverse event (AE) or other medical condition or situation occurs such that continued participation in the study would not be in the best interest of the participant
- Disease progression which requires discontinuation of the study intervention
- If the participant meets an exclusion criterion (either newly developed or not previously recognized) that precludes further study participation
- Participant unable to receive study intervention
- The reason for participant discontinuation or withdrawal from the study will be recorded on the Case Report Form (CRF). Subjects who provide informed consent and are randomized but do not receive the study intervention may be replaced. Subjects who sign the informed consent form, and are randomized and receive the study intervention, and subsequently withdraw, or are withdrawn or discontinued from the study will be replaced.
  6.7.12.3 **Lost to Follow-Up**

A participant will be considered lost to follow-up if he or she fails to return for two or more visits and is unable to be contacted by the study site staff; if a subject is hospitalized outside the YNHHS we will attempt follow for clinical outcomes and concomitant medications.

The following actions must be taken if a participant fails to return to the clinic for a required study visit:

- The site will attempt to contact the participant and reschedule the missed visit within one day and counsel the participant on the importance of maintaining the assigned visit schedule and ascertain if the participant wishes to and/or should continue in the study.
- Before a participant is deemed lost to follow-up, the investigator or designee will make every effort to regain contact with the participant (where possible, 3 telephone calls and, if necessary, a certified letter to the participant’s last known mailing address or local equivalent methods). These contact attempts should be documented in the participant’s medical record or study file.
- Should the participant continue to be unreachable, he or she will be considered to have withdrawn from the study with a primary reason of lost to follow-up.
  6.8 Study Assessments and Procedures
      6.8.1 Biological specimen collection and laboratory evaluations.

On study days 0, 2 (NP swab optional, saliva required), 4 (NP swab and saliva both required) and 6 (NP swab optional, saliva required), 14+/-2 (NP swab and saliva both required) and 28+/-2, (NP swab and saliva both required), samples will be obtained and analyzed by an FDA-authorized COVID-19 RT-PCR assay coordinated through the YNHH Clinical Virology Laboratory, and Yale Department of Pathology. Additional testing including antigen detection may be done on an NP swab, nasal swab or saliva.

On study days 0, 14+/-2 and 28+/-2, blood will be collected for 1) clinical surveillance🡪complete blood count, blood chemistries and coagulation including electrolytes, BUN, creatinine, AST, ALT, total and direct bilirubin, HS-CRP, LDH and alkaline phosphatase, PT, INR, d-dimer, fibrinogen; 2) research testing including obtaining serum, plasma and peripheral blood mononuclear cells for research studies. A maximum of 125 mL of blood will be requested to be collected on days 0, 14 and day 28 (total of 375 mL over four weeks) for these purposes.

Respiratory swab and saliva RT-PCR testing will be done by YNHHS or YNHHS-designated send-out laboratories. Quidel’s Sofia platform will be use at the study site for antigen detection. Positivity and Ct values will be recorded and log_10_ viral loads back-extrapolated from log_10_ being equivalent to 3.3 C_t_ units or as determined using a concomitant standard curve.

Blood chemistries and CBCs (and an FDA-approved POC urine pregnancy test will be done on site prior to enrollment for appropriate women as required) will be done by YNHHS laboratories and recorded, and results reported to patients.

Remaining blood samples will be stored in a biological specimen repository approved by Yale University as follows:

| **Investigator:** | Stephanie Halene |
| --- | --- |
| **Type of Review:** | Modification/Update |
| **Title of Study:** | Specimen Repository for Hematologic Diseases |
| **IRB Protocol ID:** | 1401013259 |
| **Submission ID:** | MOD00029770 |
| **Committee Name:** | IRB 0-Ad Hoc Committee |

**Final Approval Date:** 3/29/2020

**Expiration Date:** 2/17/2021

Remaining respiratory tract samples (NP swabs, saliva) processed by the YNHH Virology laboratory will be overlabelled with unique identifiers (to de-identify the samples) and conserved for future use in the Vinetz laboratory which has been approved to handle such COVID-19 specimens by EH&S

6.8.2 Safety and Other Assessments

At study visits, vital signs including pulse oximetry will be obtained. Physical examination will not be performed routinely on participants unless dictated by signs or symptoms. Any patient experiencing symptoms requiring medical examination will be referred to hospital.

On all study days, participants will be asked to answer specific questions that relate to known potential adverse effects of camostat mesylate. The previously determined incidence of adverse effects (when drug administered for at least two weeks) are:

| Symptom | 0.1 to 0.5% | <0.1% |
| --- | --- | --- |
| Rash | 0.4 |  |
| Pruritus | 0.2% |  |
| Gastrointestinal | 0.1 to 0.5% - (nausea, abdominal discomfort or fullness, diarrhea) | Anorexia, vomiting, dry mouth, heartburn, abdominal pain, constipation |

Included in the informed consent form are concerns about rare side effects. Study personnel will specifically elicit whether there are symptoms concerning for severe allergy such as jaundice, anaphylaxis or anaphylactoid reactions will be determined on a daily basis by study personnel who will ask about such symptoms (yellow eyes, weakness, wheezing, shortness of breath, tongue swelling, hives, rash) during the first 7 days of the study. Laboratory evidence of adverse effects such as thrombocytopenia, hyperbilirubinemia, or hyperkalemia will only be able to be determined at day 14.

Participants will be informed on study day 28 of all available research-specific study results to date. Results that return after day 28 will be communicated with the participant within 2 weeks of such results. If there are actionable results of routine/standard of care lab tests including CBC, chemistries and coagulation studies, they will be reported to participants in a timely manner, otherwise they will be discussed at the day 28 visit or if they become available after day 28, within 2 weeks of when new results become available.

6.9 Data Management

Electronic data capture will be used to enter study data into eCRFs and to transfer the data into a study-specific electronic database. After the study was approved, the eDC forms were reviewed by the Principal Investigator’s office, tested and validated at YCAS and YCCI. The ***Project Manager/YCAS*** prepared an Operations Manual (with assistance from the Principal Investigator) that was provided to the site investigators as a guide of the operation and management of the study as well as a technical reference manual. A training session was held at the study kick-off meeting for all study personnel in order to assure uniformity in patient management and data collection procedures, and to train the personnel in study procedures and criteria.

The study personnel at each participating center will complete and assemble the eDC forms. During the data collection process, automated quality assurance programs will be used to identify missing data, out-of-range data, and other data inconsistencies. Requests for data clarification or correction will be forwarded to the investigative study center for resolution. As appropriate, eCRFs, listings, tables, and SAS datasets will be provided to the study centers for review.

Quality assurance and quality control systems will be implemented and maintained according to written standard operating procedures to ensure that the data are generated, recorded, and reported in compliance with the protocol, GCP, and applicable regulatory requirements. Data collection and storage systems will provide audit trail, security mechanisms, and electronic signature capabilities that meet the requirements of FDA Title 21 of CFR Part 11 regarding electronic records and electronic signatures.

Data security will be controlled through appropriate and specific restriction of access only to data and systems required by individual users to accomplish their roles in the data management process. Individual login and password protections will be employed at study centers and at the sponsor or its designee. The database will exist on physically secured servers. Data backups will be done regularly and will be stored in separate facilities. Printed documents relating to the study will be secured when not under review.

6.10 Schedule of Activities

| **Procedures** | Enrollment,  Study Visit 1,  Day 0 | Study Visit 2  Day 2 | Study Visit 3  Day 4 | Study Visit 4  Day 6 | Study Visit 5  Day 14+/- 2 days | Study Visit 6  Day 28+/- 2 days |
| --- | --- | --- | --- | --- | --- | --- |
| Informed consent | X |  |  |  |  |  |
| Demographics | X |  |  |  |  |  |
| Medical history | X |  |  |  |  |  |
| Randomization | X |  |  |  |  |  |
| Administer study intervention:  Subjects receive two days worth of drug/placebo on days 0, 2, 4 and one day of drug/placebo on day 6 | X  Drug/placebo  For 2 days | X Drug/placebo  For 2 days | X  Drug/placebo  For 2 days | X Drug/placebo  For 1 day |  |  |
| Nasopharyngeal swabs, saliva for COVID-19 RT-PCR and antigen testing | X | X  Swabs optional | X | X  Swabs optional | X | X |
| Physical exam: Vital signs, wt, pulse oximetry, other as clinically indicated | X | X | X | X | X | X |
| Urine pregnancy test (as indicated) | X |  |  |  |  |  |
| Blood pregnancy test (as indicated) |  |  |  |  | X |  |
| Hematology and coagulation | X |  |  |  | X | X |
| Blood chemistries | X |  |  |  | X | X |
| Adverse event review and evaluation | X | X | X | X | X | X |
| Concomitant medication review | X | X | X | X | X | X |
| Blood for research biorespository | X |  |  |  | X | X |
| Complete Case Report Forms (CRFs) | X | X | X | X | X | X |

7. STATISTICAL CONSIDERATIONS
2. 7.1 Power and Sample Size

The sample size calculation is based on the primary outcome of interest: change in log_10_ respiratory (nasopharyngeal swab, saliva swab RT-PCR) viral load from baseline to day 4 post-randomization. Given the limited data on the variability of the change in log_10_ viral load, we have powered the study based on detecting a moderate standardized effect size of 0.3 using an analysis of covariance (ANCOVA), adjusting for baseline log_10_­ viral load. To put into context, one scenario that would produce a 0.3 standardized effect size would be a change of 4 in log_10_­ viral load in the camostat mesylate group compared to a change of 1 in log_10_­ viral load in the placebo group assuming a standard deviation of 5.0. (NOTE: For ANCOVA, the effect size is the standard deviation of the treatment means divided by the pooled standard deviations of the observations.) To be conservative we assume a R-squared of 0 between the log_10_ viral RNA at 4-days and baseline log_10_ viral RNA. With a power of 90%, and a type I error rate of 10% (2-sided), we would be able to detect the hypothesized 0.3 standardized effect size with 98 total patients - 49 patients per group with a 1:1 randomization. Increasing this sample size by 15%, 5% for an efficacy and futility look at 50% information (i.e. when half of the patients have been enrolled) and 10% to account for loss to follow up, gives a total of 114 participants (57 per treatment arm).

7.2 Interim Monitoring

Interim monitoring for safety, efficacy and futility will be conducted. When 50% of the participants have accumulated the primary endpoint, we will conduct an interim analysis to assess for efficacy and futility using the group sequential method with asymmetric boundaries. We will use a Pocock boundary for efficacy and will consider stopping early for efficacy if the t-statistic for the treatment effect is greater than the critical value of 1.875; we propose stopping for futility if the drug is trending in the wrong direction, and have set a futility boundary value of -1. Safety will be assessed by comparing the rate of adverse events in the treatment group using Type I error of 5% (2-sided); no control for multiplicity will be done for safety outcomes.

7.3 Proposed DSMB Monitoring Tables, Listings and Figures
    7.3.1 Open DSMB Report

Table 3.2 Summary of Adverse Events

7.3.2 Closed DSMB Report

8. ANALYTICAL PLAN
2. 8.1 Data Collection and Data Freeze

Data collected from Date of Study Initiation to Last Date of Study Follow-up will be used. The trial database will be closed/frozen by two months after last patient visit and be considered ready for final analysis.

8.2 General Principles

This is a double-blind placebo-controlled randomized trial testing the superiority of camostat mesylate compared to placebo for reduction of viral load. The primary comparison in the trial is between camostat and placebo. A two-sided overall type I error rate of 0.10 will be used for the primary comparison. Secondary hypotheses will be described according to the appropriate summary statistics (e.g., proportions for categorical data, means with 95% confidence intervals for continuous data). Diagnostic tests and sensitivity analyses will be performed. Parametric distributional assumptions will be checked. If assumptions fail, other distributions will be considered prior to transformations and non-parametric methods for sensitivity analysis.

This statistical analysis plan (SAP) will be finalized and filed with the study sponsor prior to database lock and unblinding.

8.3 Statistical Hypotheses

All tests performed will be tests of superiority.

For the primary endpoint, we will test the hypothesis that camostat mesylate results in a significantly greater change from baseline to day 4 in respiratory (Nasopharyngeal swab, saliva RT-PCR) log_10_ viral load compared to placebo.

Similarly, for secondary endpoints we will test the hypothesis that camostat mesylate will result in a significantly greater change from baseline to days 2 and 6 in respiratory (nasopharyngeal swab, saliva RT-PCR) log_10_ viral load compared to placebo; that camostat mesylate will reduce log_10_ of a positive COVID19 test at days 2 and 6 (optional swabs), rate of positive tests at days 6, 14 (required), and 28 (required); and that camostat mesylate will result in lower symptom burden at days 6 and 14.

8.4 Analysis Sets
    8.4.1 Intent to Treat (ITT) Population

The intention-to-treat (ITT) population includes all participants randomized according to their initial randomized assignment regardless of the treatment actually received.

8.4.2 Per Protocol (PP) Population

The Per Protocol (PP) population includes all ITT participants who did not have any major protocol deviations. The final determination of the exclusion of subjects from the PP population will be made prior to the first database lock.

8.4.3 Safety Population

The Safety population will include all randomized subjects who received at least one dose of the study drug.

8.5 Evaluation of Participant Accounting and Protocol Violations
    8.5.1 Participant Disposition

The number of patients screened (overall, by month) will be tabulated. The proportion of enrolled/eligible patients will be examined as well as reasons for exclusion. We will tabulate final participant disposition including treatment completion/discontinuation, study completion/withdrawal and lost to follow-up or death. We will summarize reasons for treatment discontinuation and study withdrawal. Study subjects who withdrew prior to study termination will be discussed in the clinical study report.

Tables, Listings and Figures (TLFs) to generate:

Figure. CONSORT diagram

Listing. Participant final off study disposition (including reasons for treatment discontinuation and study discontinuation)

Table. Summary of reasons for screen failure

Table. Summary of final disposition (i.e. treatment completion, treatment discontinuation, study completion, study withdrawal, duration on study, death)

Table. Summary of reasons for treatment discontinuation and study withdrawal

8.5.2 Protocol Violations

All major and minor known eligibility violations will be reported with a descriptive summary of each such violation. Major violations are defined as (a) errors in the process of randomization and (b) violations of inclusion/exclusion criteria, which were not reviewed and allowed/exempted.

TLFs to generate:

Listing. Protocol violations (major/minor)

8.6 Definition of Baseline and Nominal Week for measurements

For purposes of baseline summary statistics and for defining changes over time***,*** ‘Baseline’ is defined as the nearest measurement to day 0 within ***30*** days i.e. day -***30*** to randomization. If no measurement is available in this window, the first measurement after randomization BUT before the date of ***initiation of study drug*** will be used, up to day ***2***. If a patient is still missing a baseline measurement, a measurement between -***60*** and -***30*** days will be used provided the patient’s ***status*** did not change from the date of measurement to the date of randomization.

8.7 Comparability of Baseline Characteristics

In order to assess the adequacy of randomization, the distribution of baseline characteristics among the treatment groups will be compared. Distributions of baseline demographic and clinical characteristics will be summarized by intervention group. Comparability for continuous variables will be examined graphically and by summary statistics (means, medians, quartiles, etc.). Categorical variables will be examined by calculating frequency distributions. No inferential statistics will be calculated. Baseline characteristics that are determined to be unequally distributed among the treatment groups will be considered for covariate adjustment in a sensitivity analysis to determine their impact on treatment comparisons.

TLFs to generate:

Table. Summary of baseline study characteristics by treatment

8.8 Analysis of Primary Outcome: Viral Load in Nasopharyngeal Samples at Day 4

The primary analysis will be according to intent-to-treat and conducted using ANCOVA, adjusting for baseline log_10_ viral load (or conversion from Ct value assuming 1 log_10_ = 3.3 Ct units). We will use the t-statistic for the treatment effect to determine whether the treatment is statistically different from the placebo at the overall 10% level of significance, controlling for baseline log_10_ viral load. Because of the interim analysis (see below), a p-value of 0.06 (2-sided), corresponding to an efficacy boundary value of 1.875 at the last look, will be used for the level of significance for the primary outcome at the final analysis.

To make use of all available data the ANCOVA will be implemented in a repeated measures linear mixed model analysis using the following model specification:

$$Y_{ijk}=\beta_{0}+\beta_{1}time+\beta_{2}Trt*time+\varepsilon_{ijk}$$

Where:

Y*ijk* is the log_10_ viral load for the ith subject, from the jth treatment group in the kth day

*i* = 1 to 114

*j* = 1 camostat; 0 placebo

*k*= 0, 2, 4, 6

This model fits the same mean to both treatment groups at baseline (day 0) and thus conditions (i.e. adjusts) the estimates of treatment effects on baseline viral load. As viral load decreases to zero over time, we will only include timepoints up to day 6 where a sufficient number of subjects have non-zero levels.

The following sensitivity analyses will be considered:

- Compare treatment differences in the change in viral load from day 4 to baseline adjusting for age and other baseline covariates that may be different between treatment groups using the linear mixed model described above.
- Compare treatment differences in the change in viral load from day 4 to baseline using timepoints up to day 14.
- Compare treatment differences in the change in viral load from day 4 to baseline using the linear mixed model in the subgroup of participants with baseline viral load >=10^5^ copies/ml.
- Compare treatment differences in the change in viral load from day 4 to baseline using a t-test.
- Compare treatment differences in the change in viral load from day 4 to baseline using an ANCOVA with baseline viral load as a covariate and only change from baseline to day 4 as an outcome.
- Conduct a per-protocol analysis.
- Evaluate sensitivity to missing data assumption using a pattern mixture model under missing not at random assumption.

TLFs to generate:

Listing. Available outcome data for participant by time

Table. Counts and proportions of available viral load by treatment and time

Table. Reasons for missing viral load by treatment and time

Table. Raw means, sds, medians, IQR, min max by treatment and time.

Figure. Boxplot with dot overlay of log viral load by treatment and time

Figure. Plot of viral load by time for subjects

Table. LSMeans and SDs by treatment and time with t-statistic, p-value and 95% CI of difference in means

Interim Efficacy/Futility Analysis Template TLFs

**Table. Log_10_ viral load raw means by treatment group and visit.**

|  | **Camostat** | | **Placebo** | |
| --- | --- | --- | --- | --- |
| **Visit (Day)** | **N** | **Mean (SD)** | **N** | **Mean(SD)** |
| **1 (0)** | XX | X.XX (X.XX) | XX | X.XX (X.XX) |
| **2 (2)** | XX | X.XX (X.XX) | XX | X.XX (X.XX) |
| **3 (4)** | XX | X.XX (X.XX) | XX | X.XX (X.XX) |
| **4 (6)** | XX | X.XX (X.XX) | XX | X.XX (X.XX) |
| **5 (14)** | XX | X.XX (X.XX) | XX | X.XX (X.XX) |
| **6 (28)** | XX | X.XX (X.XX) | XX | X.XX (X.XX) |

**Table. Change in** l**og_10_ viral load from baseline raw means by treatment group and visit.**

|  | **Camostat** | | **Placebo** | |
| --- | --- | --- | --- | --- |
| **Visit (Day)** | **N** | **Mean (SD)** | **N** | **Mean(SD)** |
| **2 (2)** | XX | -X.XX (X.XX) | XX | -X.XX (X.XX) |
| **3 (4)** | XX | -X.XX (X.XX) | XX | -X.XX (X.XX) |
| **4 (6)** | XX | -X.XX (X.XX) | XX | -X.XX (X.XX) |
| **5 (14)** | XX | -X.XX (X.XX) | XX | -X.XX (X.XX) |
| **6 (28)** | XX | -X.XX (X.XX) | XX | -X.XX (X.XX) |

**Table. ANCOVA using linear mixed model to compare differences in viral load changes between treatment groups (n=XX).**

| **Visit (Day)** | Camostat LSMean Change (SE) | Placebo LSMean Change (SE) | Difference Placebo-Camostat (SE) | T-statistic | p-value |
| --- | --- | --- | --- | --- | --- |
| **2 (2)** | X.XX (X.XX) | X.XX (X.XX) | X.XX (X.XX) | X.XX | X.XX |
| **3 (4)** | X.XX (X.XX) | X.XX (X.XX) | X.XX (X.XX) | X.XX | X.XX |
| **4 (6)** | X.XX (X.XX) | X.XX (X.XX) | X.XX (X.XX) | X.XX | X.XX |

**Table 3.4. Sensitivity of primary outcome results to analytic model.**

| **Model** | **Analytic Method** | **N** | **Difference Between Treatment Groups (SE)** | **T-statistic** | **p-value** |
| --- | --- | --- | --- | --- | --- |
| 1 | ANCOVA Mixed Model Day 0-6 Included | XX | X.XX | X.XX | X.XX |
| 2 | ANCOVA Mixed Model Day 0 -14 Included | XX | X.XX | X.XX | X.XX |
| 3 | T-test | XX | X.XX (X.XX) | X.XX | X.XX |
| 4 | ANCOVA Day 4 Only | XX | X.XX (X.XX) | X.XX | X.XX |
| 5 | Model 1 + Adjustment for Age and Days Since Onset of Symptoms | XX | X.XX (X.XX) | X.XX | X.XX |

8.9 Analysis of Secondary Outcomes

Secondary outcomes of interest include: change in log_10_ respiratory (nasopharyngeal swab, saliva RT-PCR) viral load from baseline (day 0) to days 2 and 6 post-randomization; log_10_ and risk for a positive COVID-19 test at day 6, 14 and 28; and change of COVID-19 symptom score from baseline (day 0), as measured at days 6 and 14 +/- 2 days after enrollment (day 0). All statistical tests for secondary outcomes will be conducted at 5% level of significance (two-sided) with no adjustment for multiple testing.

8.9.1 Secondary Outcomes: Change in Viral Load From Baseline at Days 2 and 6

The same ANCOVA analysis as proposed for the primary endpoint will be used to analyze two and six day change in log_10_ for the secondary endpoint.

8.9.2 Secondary Outcomes: Risk for positive COVID19 test at day 6, 14 and 28

Risk for positive COVID19 test at day 6, 14 and 28 will be compared using an exact binomial distribution.

8.9.3 Secondary Outcome: COVID-19 PRO Daily Self-Score

The COVID-19 PRO daily self-score tool, derived almost entirely from a validated influenza symptom scoring tool (FLU-PRO) consists of 39 items that are answered daily. Items 1-33 are likert scale questions (rated 0-4) where 0 = not at all,11, 12 and 4 = very much. These items are summed to score the severity of symptoms- where a total score of 132 would indicate the greatest severity of symptoms and a score of 0 would indicate no severity of symptoms. Items 34-38 are also likert scale questions (rated 0-4) that measure the frequency of specific daily symptoms where 0 = 0 times and 4 = 4 times or more. These items are summed to score the frequency of symptoms- where the highest score for the frequency of symptoms (20) indicates the greatest burden of symptom frequency. The last question (39) asks patients for their highest temperature in Fahrenheit.

Linear mixed models will be used to describe trajectories of change in the symptom score. The models will include fixed effects for treatment, time and the interaction of treatment and time. An unstructured covariance pattern will allow for correlation between repeated observations. Differences in means at each timepoint will be estimated along with 95% confidence intervals.

8.10 Exploratory Endpoints
     8.10.1 Level of SARS-CoV-2 antigen from Nasopharyngeal specimens from baseline to days 6 and 14

TBD: There is no specific *a priori* power analysis for this exploratory analysis, however, the endpoint will provide mechanistic data regarding viral antigen levels after treatment with camostat mesylate.

8.10.2 Progression to Hospitalization or Other Severe Disease

TBD: To gain insight into whether clinical outcomes might improve after treatment with camostat mesylate 200 mg po QID but this study is not powered to assess.

8.11 Safety and Tolerability Analyses

Adverse events (AEs) and serious adverse events (SAEs) will be categorized by severity and relation to study drug using the Safety population. AEs will be classified according to the National Cancer Institute Common Terminology Criteria for Adverse Events, version 5.0.

Safety endpoints including SAEs, Grade 3 and 4 adverse events and signs compatible with cardiac arrhythmias, allergic reactions, gastrointestinal, hepatobiliary, eye and ear, blood and lymphatic system disorders and acute exacerbation of lung disorders will be described by treatment group. Each AE will be counted once for a given participant and graded by severity and relationship to COVID-19 or study intervention. AEs will be presented by system organ class, duration (in days), start- and stop-date. Adverse events leading to premature discontinuation from the study intervention and serious treatment-emergent AEs will be presented in tables and listings.

Safety will be reported by treatment arm using descriptive statistics for laboratory assessments (numeric summaries of change from baseline and shift tables) and counts/proportions/rates for adverse events. A type I error of 5% (2-sided) will be used with no control for multiplicity. We will consider Wilcoxon rank sum tests or two-sample t-tests for continuous outcomes and the exact binomial distribution for proportions.

8.12 Adverse Events

Treatment-emergent AEs and non-TEAEs (those occurring prior to administration of study medication or that first occurred prior to study drug administration and did not worsen in frequency or severity) will be listed. TEAEs will be defined as AEs that occur on or after the date and time of study drug administration, or those that first occur pre-dose but worsen in frequency or severity after study drug administration. AEs will be followed-up until complete resolution, or until the PI or Sub-Investigator judges safe to discontinue follow-up.

The incidence of TEAEs will be summarized using the safety population. The MedDRA® dictionary Version 23 will be used to classify all AEs reported during the study by system organ class (SOC) and preferred term (PT). Incidence of subjects who experienced TEAEs will be presented by treatment and overall, SOC, PT, by Investigator-assessed relationship and also by severity. Each subject may only contribute once to each of the incidence rates, for a TEAE following a given treatment, regardless of the number of occurrences; the highest severity or highest relationship will be presented, as appropriate. In each table, SOC will be presented in descending order of overall incidence rate in terms of frequency of subjects and then in frequency of events (alphabetical order will be used in case of equal rates). For each SOC, PT will be presented the same way.

Incidence of TEAEs (number of events) will also be presented by treatment and overall, by SOC, and PT, by Investigator-assessed relationship and severity.

Severity of AEs will be rated according to CTCAE criteria as outlined in Section 6.7 and defined in Table 6.7.1 as Grade 1 (mild), Grade 2 (moderate), Grade 3 (severe), Grade 4 (life-threatening) and Grade 5 (fatal).

TLFs to generate:

Listing. Participant deaths, description and follow-up time

Listing. Participant SAEs, grade, SOC, PT, onset, resolve date, relation to treatment, action taken

Listing. Participant AEs, grade, SOC, PT, onset, resolve date, relation to treatment

Table. Counts, person months, rates and proportions of Death and SAEs by treatment group

Table. Counts, person months, rates and proportions of AEs by treatment group

Table. Counts, person months, rates and proportions of AEs by treatment group by grade

8.13 Laboratory Parameters

Clinical laboratory tests including biochemistry, hematology, and urinalysis, and other screening tests, will be performed at time points specified in the SoA (Section 6.10). Microscopic examination will be performed on abnormal findings. Listings of all clinical laboratory results will be provided with the abnormal values flagged with "L" (Below normal range) and "H" (Above normal range) for continuous parameters, and “N” (Normal Range) for categorical parameters.

Descriptive statistics (mean, median, SD, Min, Max, and sample size) for each clinical laboratory test (continuous variables) will be presented. Change from baseline to study exit will also be presented. For a given laboratory test, any value (scheduled or unscheduled), prior to the first dosing will replace any missing screening value (if scheduled value is missing). However, unscheduled results will not be included in the summary tables. For categorical variable (urinalysis test), the number of subjects (frequency and percentage) will be tabulated by results (e.g., negative, positive, trace, etc.). A summary table of shifts from screening to study exit will be provided. Results from repeat tests will not be included in the summary statistics unless the repeat was required (and documented as such) due to technical reasons or an invalid initial result.

TLFs to generate:

Listing. Abnormal laboratory results

Table. Laboratory Results Summary - baseline, each visit, change from baseline - mean (sd), median (min, max)

Tables. Shift tables for baseline to follow-up

8.14 Plan for Missing Data

Several strategies will be imposed to minimize the impact of missing data on conclusions. Prevention is the most obvious and effective manner to control bias and loss of power from missing data (NRC 2010). This protocol will follow the intent to treat principle, requiring follow-up of all subjects randomized regardless of the actual treatment received (Lachin, 2000). The only reasons for study withdrawal will be withdrawal of consent and lost to follow-up. The informed consent and protocol will distinguish between treatment discontinuation and study withdrawal and the consent will also include a statement about the importance of continuing data collection despite treatment discontinuation. Alternative contact information will be identified on entry into the study to minimize loss-to follow-up. Timely data entry combined with weekly missing data reports will trigger protocols for tracking and obtaining missing data items or outcome assessments. Despite these prevention efforts it is reasonable to assume missing data will occur. The primary analysis is valid under the assumption that missing data is missing at random (MAR). Logistic regression will be used to identify factors associated with dropout. While differential rates of dropout between groups or high loss to follow-up are not expected, sensitivity analysis using pattern-mixture and selection models under missing not at random (MNAR) assumptions will be performed to examine the robustness of conclusions of the primary analysis to missing data.

8.15 Subgroup Analysis

Heterogeneity of treatment effect (HTE) for the primary and secondary progression to hospitalization or death outcome will be examined. HTE will be evaluated within a repeated measures mixed model for the primary outcome and a logistic regression for the secondary progression outcome. The models will include treatment, site, the subgroup variable, and the interaction of the subgroup variable with time. Differences in means or risk with 95% confidence intervals will be estimated for the treatment difference within each level of the subgroup variable. The significance test of the interaction will be used to formally test HTE at the two-sided 0.05 significance level.

Pre-specified subgroup analyses will be conducted for the following:

TBD

TLFs to generate:

Table. Summary of Day 4 viral load by treatment and subgroup level

Figure. Forest plot of risk difference with 95% CI by subgroups

APPENDIX

APPENDIX 1. OPEN REPORT TEMPLATE TABLES, LISTING, FIGURES

Figure 1.1. Cumulative Number Enrolled by Month

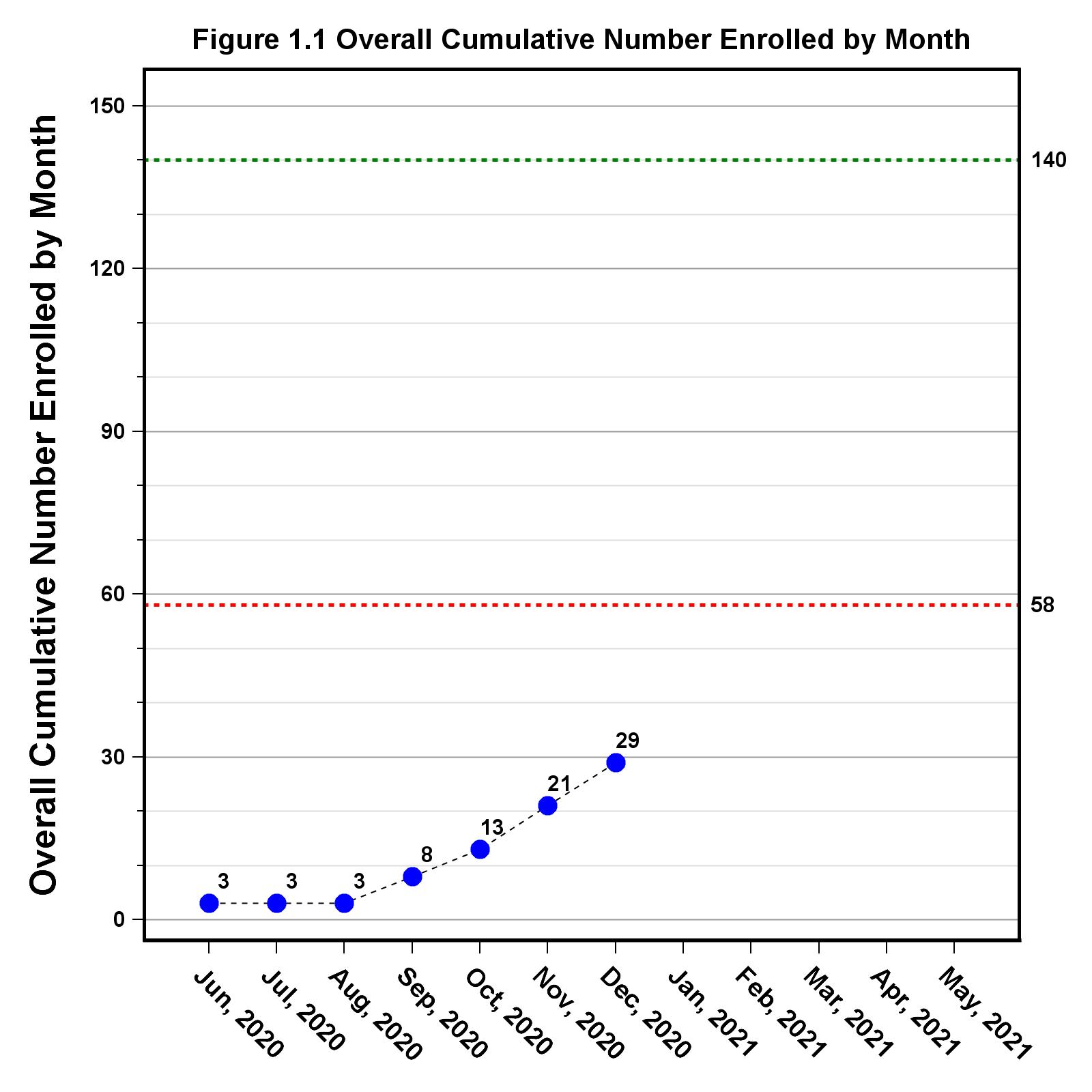

Table 1.1. Participant Disposition

|  | ***Total N (%)*** |
| --- | --- |
| **Randomized** | XX (XXX.X%) |
| **On Study** | X ( X.X%) |
| On treatment | X ( X.X%) |
| Off treatment | X ( X.X%) |
| **Off Study** | XX (XXX.X%) |
| Study Completed | XX ( XX.X%) |
| Withdrawal | X ( XX.X%) |
| Lost Follow-up | X ( X.X%) |
| Death | X ( X.X%) |
| Other | X ( X.X%) |
| **Treatment Disposition** | XX |
| On Treatment | X ( X.X%) |
| Off Treatment | XX (XXX.X%) |
| Study Completed | XX ( XX.X%) |
| PI decision due to AE | X ( X.X%) |
| PI decision due to non-compliance | X ( X.X%) |
| Hospitalization | X ( X.X%) |
| Subject declined due to AE | X ( X.X%) |
| Subject declined due to feeling better | X ( X.X%) |
| Other | X ( X.X%) |

Table 2.1. Baseline Characteristics

|  | ***Total (N=XX)*** |
| --- | --- |
| **Age (years)** | |
| N | XX |
| Mean (SD) | XX.XX (XX.XX) |
| Median | XX.XX |
| Min, Max | XX.XX, XX.XX |
| **Sex** | |
| Male | XX (XX.X%) |
| Female | XX (XX.X%) |
| **Race** | |
| White | XX (XX.X%) |
| Black or African American | X ( X.X%) |
| Asian | X ( X.X%) |
| Native Hawaiian or Other Pacific Islander | X ( X.X%) |
| American Indian or Alaska Native | X ( X.X%) |
| Other | X ( X.X%) |
| Unknown | X ( X.X%) |
| **Ethnicity** | |
| Hispanic or Latino | X (XX.X%) |
| Not Hispanic or Latino | XX (XX.X%) |
| Not reported | X ( X.X%) |
| Unknown | X ( X.X%) |

Table 2.2 Completed Study Visits

| **Visit** | **Total Expected Visits** | **Completed Visits** | **% Completed Visits** | **Not Completed Visits** | **% Not Completed Visits** |
| --- | --- | --- | --- | --- | --- |
| Screening | XX | XX | XXX.X% | X | X.X% |
| Visit 1 (Day 0) | XX | XX | XXX.X% | X | X.X% |
| Visit 2 (Day 2) | XX | XX | XXX.X% | X | X.X% |
| Visit 3 (Day 4) | XX | XX | XX.X% | X | X.X% |
| Visit 4 (Day 6) | XX | XX | XX.X% | X | X.X% |
| Visit 5 (Day 14) | XX | XX | XXX.X% | X | X.X% |
| Visit 6 (Day 28) | XX | XX | XXX.X% | X | X.X% |
| Visit Totals | XXX | XXX | XX.X% | X | X.X% |

Table 2.3 Completed CRFs by Visit

| **Visit** | **Total Expected CRFs** | **Completed CRFs** | **% Completed CRFs** | **Not Completed CRFs** | **% Not Completed CRFs** | **Missed CRFs** | **% Missed CRFs** |
| --- | --- | --- | --- | --- | --- | --- | --- |
| Screening | XX | XX | XXX.X% | X | X.X% | X | X.X% |
| Visit 1 (Day 0) | XXX | XXX | XXX.X% | X | X.X% | X | X.X% |
| Visit 2 (Day 2) | XXX | XXX | XX.X% | X | X.X% | X | X.X% |
| Visit 3 (Day 4) | XXX | XXX | XX.X% | X | X.X% | X | X.X% |
| Visit 4 (Day 6) | XXX | XX | XX.X% | X | X.X% | X | X.X% |
| Visit 5 (Day 14) | XXX | XXX | XX.X% | X | X.X% | X | X.X% |
| Visit 6 (Day 28) | XXX | XXX | XX.X% | X | X.X% | X | X.X% |
| Visit Totals | XXX | XXX | XX.X% | X | X.X% | XX | X.X% |

Table 2.4 Completed COVID-19 Patient Reported Outcomes Forms

| **Visit** | **Total Expected CRFs** | **Completed CRFs** | **% Completed CRFs** | **Not Completed CRFs** | **% Not Completed CRFs** | **Missed CRFs** | **% Missed CRFs** |
| --- | --- | --- | --- | --- | --- | --- | --- |
| Day 1 | XX | XX | XXX.X% | X | X.X% | X | X.X% |
| Day 3 | XX | XX | XXX.X% | X | X.X% | X | X.X% |
| Day 5 | XX | XX | XXX.X% | X | X.X% | X | X.X% |
| Day 7 | XX | XX | XX.X% | X | XX.X% | X | X.X% |
| Day 8 | XX | XX | XX.X% | X | XX.X% | X | X.X% |
| Day 9 | XX | XX | XX.X% | X | XX.X% | X | X.X% |
| Day 10 | XX | XX | XX.X% | X | XX.X% | X | X.X% |
| Day 11 | XX | XX | XX.X% | X | XX.X% | X | X.X% |
| Day 12 | XX | XX | XX.X% | X | XX.X% | X | X.X% |
| Day 13 | XX | XX | XX.X% | X | XX.X% | X | X.X% |
| Visit Totals | XXX | XXX | XX.X% | XX | XX.X% | X | X.X% |

Table 3.1 Summary of Serious Adverse Events

|  | ***Total (N=XX)*** |
| --- | --- |
| **Total Number of Serious Adverse Events** | X |
| **Number of Participants With at Least 1 Adverse Event** | X ( X.X%) |
| **Respiratory, thoracic and mediastinal disorders** | X (XXX.X%) |
| Dyspnea | X ( XX.X%) |
| Respiratory, thoracic and mediastinal disorders – Other, specify | X ( XX.X%) |
| ….. | |
| ….. | |

Table 3.2 Summary of Adverse Events

|  | ***Total (N=XX)*** |
| --- | --- |
| **Total Number of Adverse Events** | XX |
| **Number of Participants With at Least One Adverse Event** | X ( XX.X%) |
| Respiratory, Thoracic And Mediastinal Disorders | X ( XX.X%) |
| Gastrointestinal Disorders | X ( XX.X%) |
| Nervous System Disorders | X ( XX.X%) |
| Blood And Lymphatic System Disorders | X ( XX.X%) |
| Cardiac Disorders | X ( XX.X%) |
| Investigations | X ( XX.X%) |
| …..  ….. | |

Listing 3.1 Serious Adverse Events

| ***Subject*** | ***System Organ Class/ Preferred Term/ Verbatim**** | ***Start Date/ End Date*** | ***Outcome*** | ***Action Taken with Study Treatment*** | ***Relationship to Study Treatment*** | ***Serious*** | ***Life Threatening*** | ***Hospitalization*** | ***Death*** | ***Significant Disability*** | ***Congenital Anomaly or Birth Defect*** | ***Toxicity Grade*** |
| --- | --- | --- | --- | --- | --- | --- | --- | --- | --- | --- | --- | --- |
| XXXXX | XXXXXXXXXXXXXXXXXXXXXXXXXXXXXXXXXXXXXXXXXXXXXXXXXXXXXXXXXXXXXXXXXXXXXXXXXXXXX | XXXXXXXXXXXXXXXXXXXX | XXXXXXXXXXXXXXXXXXX | XXXXXXXXXXXXXXXX | XXXXXXXX | XXX | XXX | XXX | XX | XX | XX | X |
| XXXXX | XXXXXXXXXXXXXXXXXXXXXXXXXXXXXXXXXXXXXXXXXXXXXXXXXXXXXXXXXXXXXXXXXXXXXXXXXXXXXXXXXXXXXXXXXXXXXXXXXXXXXXXXXXXXXXXXXXXXXXXXXXXXXXXXXXXXXXXX | XXXXXXXXXXXXXXXXXXXX | XXXXXXXXXXXXXXXXXXX | XXXXXXXXXXXXXX | XXXXXXXX | XXX | XX | XXX | XX | XX | XX | X |

Listing 3.2 Adverse Events

| ***Subject*** | ***Early Termination*** | ***System Organ Class/ Preferred Term/ Verbatim**** | ***Start Date/ End Date*** | ***Outcome*** | ***Action Taken with Study Treatment*** | ***Relationship to Study Treatment*** | ***Serious*** | ***Life Threatening*** | ***Hospitalization*** | ***Death*** | ***Significant Disability*** | ***Congenital Anomaly or Birth Defect*** | ***Toxicity Grade*** |
| --- | --- | --- | --- | --- | --- | --- | --- | --- | --- | --- | --- | --- | --- |
| XXXXX | XX | XXXXXXXXXXXXXXXXXXXXXXXXXXXXXXXXXXXXXXXXXXXXXXXXXXXXXXXXXXXXXXXXXXXXXXXXXXXXXXXXXXXXXXXXXXXXXXXXX | XXXXXXXXXXXXXXXXXXXX | XXXXXXXXXXXXXXXXXXX | XXXX | XXXXXXXXX | XX |  |  |  |  |  | X |

Table A3.1 Summary of Serious Adverse Events by Relationship to Study Drug

|  | ***Total (N=XX)*** | | | | |
| --- | --- | --- | --- | --- | --- |
|  | ***Unrelated*** | ***Unlikely*** | ***Possible*** | ***Probable*** | ***Definite*** |
| **Total Number of Adverse Events** | X | X | X | X | X |
| **Number of Participants With at Least 1 Adverse Event** | X ( X.X%) | X ( X.X%) | X ( X.X%) | X ( X.X%) | X ( X.X%) |
| **Respiratory, thoracic and mediastinal disorders** | X ( X.X%) | X (XXX.X%) | X ( X.X%) | X ( X.X%) | X ( X.X%) |
| Dyspnea | X ( X.X%) | X ( XX.X%) | X ( X.X%) | X ( X.X%) | X ( X.X%) |
| Respiratory, thoracic and mediastinal disorders – Other, specify | X ( X.X%) | X ( XX.X%) | X ( X.X%) | X ( X.X%) | X ( X.X%) |

Table A3.2 Summary of Adverse Events by Relationship to Study Drug

|  | ***Total (N=XX)*** | | | | |
| --- | --- | --- | --- | --- | --- |
|  | ***Unrelated*** | ***Unlikely*** | ***Possible*** | ***Probable*** | ***Definite*** |
| **Total Number of Adverse Events** | X | 5 | 0 | 0 | 0 |
| **Number of Participants With at Least 1 Adverse Event** | X ( XX.X%) | 4 ( 13.8%) | 0 ( 0.0%) | 0 ( 0.0%) | 0 ( 0.0%) |
| **Blood and lymphatic system disorders** | X ( X.X%) | 1 ( 20.0%) | 0 ( 0.0%) | 0 ( 0.0%) | 0 ( 0.0%) |
| Blood and lymphatic system disorders – Other, specify | X ( X.X%) | 1 ( 20.0%) | 0 ( 0.0%) | 0 ( 0.0%) | 0 ( 0.0%) |
| **Cardiac disorders** | X ( XX.X%) | 0 ( 0.0%) | 0 ( 0.0%) | 0 ( 0.0%) | 0 ( 0.0%) |
| Cardiac disorders – Other, specify | X ( XX.X%) | 0 ( 0.0%) | 0 ( 0.0%) | 0 ( 0.0%) | 0 ( 0.0%) |
| **Gastrointestinal disorders** | X ( X.X%) | 2 ( 40.0%) | 0 ( 0.0%) | 0 ( 0.0%) | 0 ( 0.0%) |
| Diarrhea | X ( X.X%) | 1 ( 20.0%) | 0 ( 0.0%) | 0 ( 0.0%) | 0 ( 0.0%) |
| Gastroesophageal reflux disease | X ( X.X%) | 1 ( 20.0%) | 0 ( 0.0%) | 0 ( 0.0%) | 0 ( 0.0%) |
| **Investigations** | X ( XX.X%) | 0 ( 0.0%) | 0 ( 0.0%) | 0 ( 0.0%) | 0 ( 0.0%) |
| Investigations – Other, specify | X ( XX.X%) | 0 ( 0.0%) | 0 ( 0.0%) | 0 ( 0.0%) | 0 ( 0.0%) |
| **Nervous system disorders** | X ( XX.X%) | 0 ( 0.0%) | 0 ( 0.0%) | 0 ( 0.0%) | 0 ( 0.0%) |
| Presyncope | X ( XX.X%) | 0 ( 0.0%) | 0 ( 0.0%) | 0 ( 0.0%) | 0 ( 0.0%) |
| Syncope | X ( XX.X%) | 0 ( 0.0%) | 0 ( 0.0%) | 0 ( 0.0%) | 0 ( 0.0%) |
| **Respiratory, thoracic and mediastinal disorders** | X ( XX.X%) | 2 ( 40.0%) | 0 ( 0.0%) | 0 ( 0.0%) | 0 ( 0.0%) |
| Dyspnea | X ( X.X%) | 1 ( 20.0%) | 0 ( 0.0%) | 0 ( 0.0%) | 0 ( 0.0%) |
| Respiratory, thoracic and mediastinal disorders - Other, specify | X ( X.X%) | 1 ( 20.0%) | 0 ( 0.0%) | 0 ( 0.0%) | 0 ( 0.0%) |
| Sore throat  …… | X ( XX.X%) | 0 ( 0.0%) | 0 ( 0.0%) | 0 ( 0.0%) | 0 ( 0.0%) |

Table A3.3 Summary of Serious Adverse Events by Severity

|  | ***Total (N=XX)*** | | |
| --- | --- | --- | --- |
|  | ***Severe*** | ***Life Threatening*** | ***Death*** |
| **Total Number of Adverse Events** | X | X | X |
| **Number of Participants With at Least X Adverse Event** | X ( X.X%) | X ( X.X%) | X ( X.X%) |
| **Respiratory, thoracic and mediastinal disorders** | X (XXX.X%) | X (XXX.X%) | X ( X.X%) |
| Dyspnea | X ( X.X%) | X (XXX.X%) | X ( X.X%) |
| Respiratory, thoracic and mediastinal disorders - Other, specify  …… | X (XXX.X%) | X ( X.X%) | X ( X.X%) |

Table A3.4 Summary of Adverse Events by Severity

|  | ***Total (N=XX)*** | | | |
| --- | --- | --- | --- | --- |
|  | ***Mild Moderate*** | ***Severe*** | ***Life Threatening*** | ***Death*** |
| **Total Number of Adverse Events** | X | X | X | X |
| **Number of Participants With at Least 1 Adverse Event** | X ( XX.X%) | X ( X.X%) | X ( X.X%) | X ( X.X%) |
| **Blood and lymphatic system disorders** | X ( XX.X%) | X ( X.X%) | X ( X.X%) | X ( X.X%) |
| Blood and lymphatic system disorders - Other, specify | X ( XX.X%) | X ( X.X%) | X ( X.X%) | X ( X.X%) |
| **Cardiac disorders** | X ( XX.X%) | X ( X.X%) | X ( X.X%) | X ( X.X%) |
| Cardiac disorders - Other, specify | X ( XX.X%) | X ( X.X%) | X ( X.X%) | X ( X.X%) |
| **Gastrointestinal disorders** | X ( XX.X%) | X ( X.X%) | X ( X.X%) | X ( X.X%) |
| Diarrhea | X ( XX.X%) | X ( X.X%) | X ( X.X%) | X ( X.X%) |
| Gastroesophageal reflux disease | X ( XX.X%) | X ( X.X%) | X ( X.X%) | X ( X.X%) |
| **Investigations** | X ( XX.X%) | X ( X.X%) | X ( X.X%) | X ( X.X%) |
| Investigations - Other, specify | X ( XX.X%) | X ( X.X%) | X ( X.X%) | X ( X.X%) |
| **Nervous system disorders** | X ( XX.X%) | X ( X.X%) | X ( X.X%) | X ( X.X%) |
| Presyncope | X ( XX.X%) | X ( X.X%) | X ( X.X%) | X ( X.X%) |
| Syncope | X ( XX.X%) | X ( X.X%) | X ( X.X%) | X ( X.X%) |
| **Respiratory, thoracic and mediastinal disorders** | X ( XX.X%) | X (XXX.X%) | X (XXX.X%) | X ( X.X%) |
| Dyspnea | X ( X.X%) | X ( X.X%) | X (XXX.X%) | X ( X.X%) |
| Respiratory, thoracic and mediastinal disorders - Other, specify | X ( X.X%) | X (XXX.X%) | X ( X.X%) | X ( X.X%) |
| Sore throat | X ( XX.X%) | X ( X.X%) | X ( X.X%) | X ( X.X%) |

Table A4.1 Summary of Lab Changes – Hemoglobin

|  | ***Total (N=XX)*** | |
| --- | --- | --- |
|  | ***Absolute Result*** | ***Change From Baseline*** |
| **Hemoglobin (g/dL)** | | |
| **Baseline (Visit 1/Day 0)** | | |
| N | XX |  |
| Mean (SD) | XX.X (X.X) |  |
| Median | XX.X |  |
| Min, Max | XX.X, XX.X |  |
| **Visit 5/Day 14** | | |
| N | XX | XX |
| Mean (SD) | XX.X (X.X) | -X.X (X.X) |
| Median | XX.X | -X.X |
| Min, Max | XX.X, XX.X | -XX.X, X.X |
| **Visit 6/Day 28** | | |
| N | XX | XX |
| Mean (SD) | XX.X (X.X) | X.X (XX.X) |
| Median | XX.X | -X.X |
| Min, Max | XX.X, XX.X | -XX.X, XX.X |

Table A4.2 Summary of Lab Changes – Hematocrit

|  | ***Total (N=XX)*** | |
| --- | --- | --- |
|  | ***Absolute Result*** | ***Change From Baseline*** |
| **Hematocrit (%)** | | |
| **Baseline (Visit 1/Day 0)** | | |
| N | XX |  |
| Mean (SD) | XX.X (X.X) |  |
| Median | XX.X |  |
| Min, Max | XX.X, XX.X |  |
| **Visit 5/Day 14** | | |
| N | XX | XX |
| Mean (SD) | XX.X (X.X) | -X.X (X.X) |
| Median | XX.X | -X.X |
| Min, Max | XX.X, XX.X | -X.X, XX.X |
| **Visit 6/Day 28** | | |
| N | XX | XX |
| Mean (SD) | XX.X (X.X) | -X.X (XX.X) |
| Median | XX.X | -X.X |
| Min, Max | XX.X, XX.X | -XX.X, XX.X |

Table A4.3 Summary of Lab Changes – WBC

|  | ***Total (N=XX)*** | |
| --- | --- | --- |
|  | ***Absolute Result*** | ***Change From Baseline*** |
| **WBC (x1000/uL)** | | |
| **Baseline (Visit 1/Day 0)** | | |
| N | XX |  |
| Mean (SD) | X.X (X.X) |  |
| Median | X.X |  |
| Min, Max | X.X, XX.X |  |
| **Visit 5/Day 14** | | |
| N | XX | XX |
| Mean (SD) | X.X (X.X) | X.X (X.X) |
| Median | X.X | X.X |
| Min, Max | X.X, XX.X | -X.X, X.X |
| **Visit 6/Day 28** | | |
| N | XX | XX |
| Mean (SD) | X.X (X.X) | X.X (X.X) |
| Median | X.X | X.X |
| Min, Max | X.X, X.X | -X.X, X.X |

Table A4.4 Summary of Lab Changes – ALC

|  | ***Total (N=XX)*** | |
| --- | --- | --- |
|  | ***Absolute Result*** | ***Change From Baseline*** |
| **ALC (x1000/uL)** | | |
| **Baseline (Visit 1/Day 0)** | | |
| N | XX |  |
| Mean (SD) | X.X (XX.X) |  |
| Median | X.X |  |
| Min, Max | X.X, XXX.X |  |
| **Visit 5/Day 14** | | |
| N | XX | XX |
| Mean (SD) | X.X (X.X) | -X.X (XX.X) |
| Median | X.X | X.X |
| Min, Max | X.X, X.X | -XXX.X, X.X |
| **Visit 6/Day 28** | | |
| N | XX | XX |
| Mean (SD) | X.X (X.X) | -X.X (XX.X) |
| Median | X.X | X.X |
| Min, Max | X.X, X.X | -XXX.X, X.X |

Table A4.5 Summary of Lab Changes – Platelet Count

|  | ***Total (N=XX)*** | |
| --- | --- | --- |
|  | ***Absolute Result*** | ***Change From Baseline*** |
| **Platelet Count (x1000/uL)** | | |
| **Baseline (Visit 1/Day 0)** | | |
| N | XX |  |
| Mean (SD) | XXX.X (XX.X) |  |
| Median | XXX.X |  |
| Min, Max | X.X, XXX.X |  |
| **Visit 5/Day 14** | | |
| N | XX | XX |
| Mean (SD) | XXX.X (XX.X) | XX.X (XX.X) |
| Median | XXX.X | XX.X |
| Min, Max | XXX.X, XXX.X | -XX.X, XXX.X |
| **Visit 6/Day 28** | | |
| N | XX | XX |
| Mean (SD) | XXX.X (XX.X) | XX.X (XX.X) |
| Median | XXX.X | XX.X |
| Min, Max | XXX.X, XXX.X | -XXX.X, XXX.X |

Table A4.6 Summary of Lab Changes – Sodium

|  | ***Total (N=XX)*** | |
| --- | --- | --- |
|  | ***Absolute Result*** | ***Change From Baseline*** |
| **Sodium (mmol/l)** | | |
| **Baseline (Visit 1/Day 0)** | | |
| N | XX |  |
| Mean (SD) | XXX.X (X.X) |  |
| Median | XXX.X |  |
| Min, Max | XXX.X, XXX.X |  |
| **Visit 5/Day 14** | | |
| N | XX | XX |
| Mean (SD) | XXX.X (X.X) | X.X (X.X) |
| Median | XXX.X | X.X |
| Min, Max | XXX.X, XXX.X | -X.X, XX.X |
| **Visit 6/Day 28** | | |
| N | XX | XX |
| Mean (SD) | XXX.X (XX.X) | -X.X (XX.X) |
| Median | XXX.X | -X.X |
| Min, Max | XX.X, XXX.X | -XXX.X, XX.X |

Table A4.7 Summary of Lab Changes – Potassium

|  | ***Total (N=XX)*** | |
| --- | --- | --- |
|  | ***Absolute Result*** | ***Change From Baseline*** |
| **Potassium (mmol/l)** | | |
| **Baseline (Visit 1/Day 0)** | | |
| N | XX |  |
| Mean (SD) | X.X (X.X) |  |
| Median | X.X |  |
| Min, Max | X.X, X.X |  |
| **Visit 5/Day 14** | | |
| N | XX | XX |
| Mean (SD) | X.X (X.X) | X.X (X.X) |
| Median | X.X | X.X |
| Min, Max | X.X, X.X | -X.X, X.X |
| **Visit 6/Day 28** | | |
| N | XX | XX |
| Mean (SD) | X.X (X.X) | X.X (X.X) |
| Median | X.X | -X.X |
| Min, Max | X.X, X.X | -X.X, X.X |

Table A4.8 Summary of Lab Changes – BUN

|  | ***Total (N=XX)*** | |
| --- | --- | --- |
|  | ***Absolute Result*** | ***Change From Baseline*** |
| **BUN (mg/dL)** | | |
| **Baseline (Visit 1/Day 0)** | | |
| N | XX |  |
| Mean (SD) | XX.X (X.X) |  |
| Median | XX.X |  |
| Min, Max | X.X, XX.X |  |
| **Visit 5/Day 14** | | |
| N | XX | XX |
| Mean (SD) | XX.X (X.X) | -X.X (X.X) |
| Median | XX.X | X.X |
| Min, Max | X.X, XX.X | -X.X, X.X |
| **Visit 6/Day 28** | | |
| N | XX | XX |
| Mean (SD) | XX.X (X.X) | X.X (X.X) |
| Median | XX.X | X.X |
| Min, Max | X.X, XX.X | -XX.X, X.X |

Table A4.9 Summary of Lab Changes – Creatinine

|  | ***Total (N=XX)*** | |
| --- | --- | --- |
|  | ***Absolute Result*** | ***Change From Baseline*** |
| **Creatinine (mg/dL)** | | |
| **Baseline (Visit 1/Day 0)** | | |
| N | XX |  |
| Mean (SD) | X.X (X.X) |  |
| Median | X.X |  |
| Min, Max | X.X, X.X |  |
| **Visit 5/Day 14** | | |
| N | XX | XX |
| Mean (SD) | X.X (X.X) | -X.X (X.X) |
| Median | X.X | -X.X |
| Min, Max | X.X, X.X | -X.X, X.X |
| **Visit 6/Day 28** | | |
| N | XX | XX |
| Mean (SD) | X.X (X.X) | X.X (X.X) |
| Median | X.X | -X.X |
| Min, Max | X.X, X.X | -X.X, X.X |

Table A4.10 Summary of Lab Changes – AST

|  | ***Total (N=XX)*** | |
| --- | --- | --- |
|  | ***Absolute Result*** | ***Change From Baseline*** |
| **AST (U/L)** | | |
| **Baseline (Visit 1/Day 0)** | | |
| N | XX |  |
| Mean (SD) | XX.X (X.X) |  |
| Median | XX.X |  |
| Min, Max | XX.X, XX.X |  |
| **Visit 5/Day 14** | | |
| N | XX | XX |
| Mean (SD) | XX.X (X.X) | -X.X (X.X) |
| Median | XX.X | -X.X |
| Min, Max | XX.X, XX.X | -XX.X, XX.X |
| **Visit 6/Day 28** | | |
| N | XX | XX |
| Mean (SD) | XX.X (XX.X) | -X.X (X.X) |
| Median | XX.X | -X.X |
| Min, Max | XX.X, XX.X | -XX.X, XX.X |

Table A4.11 Summary of Lab Changes – ALT

|  | ***Total (N=XX)*** | |
| --- | --- | --- |
|  | ***Absolute Result*** | ***Change From Baseline*** |
| **ALT (U/L)** | | |
| **Baseline (Visit 1/Day 0)** | | |
| N | XX |  |
| Mean (SD) | XX.X (XX.X) |  |
| Median | XX.X |  |
| Min, Max | XX.X, XXX.X |  |
| **Visit 5/Day 14** | | |
| N | XX | XX |
| Mean (SD) | XX.X (XX.X) | X.X (XX.X) |
| Median | XX.X | X.X |
| Min, Max | XX.X, XXX.X | -XX.X, XX.X |
| **Visit 6/Day 28** | | |
| N | XX | XX |
| Mean (SD) | XX.X (XX.X) | -X.X (XX.X) |
| Median | XX.X | -X.X |
| Min, Max | X.X, XX.X | -XX.X, XX.X |

Table A4.12 Summary of Lab Changes – Total Bilirubin

|  | ***Total (N=XX)*** | |
| --- | --- | --- |
|  | ***Absolute Result*** | ***Change From Baseline*** |
| **Total Bilirubin (mg/dL)** | | |
| **Baseline (Visit 1/Day 0)** | | |
| N | XX |  |
| Mean (SD) | X.X (X.X) |  |
| Median | X.X |  |
| Min, Max | X.X, X.X |  |
| **Visit 5/Day 14** | | |
| N | XX | XX |
| Mean (SD) | X.X (X.X) | X.X (X.X) |
| Median | X.X | X.X |
| Min, Max | X.X, X.X | -X.X, X.X |
| **Visit 6/Day 28** | | |
| N | XX | XX |
| Mean (SD) | X.X (X.X) | X.X (X.X) |
| Median | X.X | X.X |
| Min, Max | X.X, X.X | -X.X, X.X |

Table A4.13 Summary of Lab Changes – Alkaline Phosphatase

|  | ***Total (N=XX)*** | |
| --- | --- | --- |
|  | ***Absolute Result*** | ***Change From Baseline*** |
| **Alkaline Phosphatase (U/L)** | | |
| **Baseline (Visit 1/Day 0)** | | |
| N | XX |  |
| Mean (SD) | XX.X (XX.X) |  |
| Median | XX.X |  |
| Min, Max | XX.X, XXX.X |  |
| **Visit 5/Day 14** | | |
| N | XX | XX |
| Mean (SD) | XX.X (XX.X) | -X.X (XX.X) |
| Median | XX.X | X.X |
| Min, Max | XX.X, XXX.X | -XX.X, XX.X |
| **Visit 6/Day 28** | | |
| N | XX | XX |
| Mean (SD) | XX.X (XX.X) | -X.X (X.X) |
| Median | XX.X | -X.X |
| Min, Max | XX.X, XXX.X | -XX.X, XX.X |

Table A4.14 Summary of Lab Changes – LDH

|  | ***Total (N=XX)*** | |
| --- | --- | --- |
|  | ***Absolute Result*** | ***Change From Baseline*** |
| **LDH (U/L)** | | |
| **Baseline (Visit 1/Day 0)** | | |
| N | XX |  |
| Mean (SD) | XXX.X (XX.X) |  |
| Median | XXX.X |  |
| Min, Max | XXX.X, XXX.X |  |
| **Visit 5/Day 14** | | |
| N | XX | XX |
| Mean (SD) | XXX.X (XX.X) | X.X (XX.X) |
| Median | XXX.X | -X.X |
| Min, Max | XXX.X, XXX.X | -XX.X, XXX.X |
| **Visit 6/Day 28** | | |
| N | XX | XX |
| Mean (SD) | XXX.X (XX.X) | X.X (XX.X) |
| Median | XXX.X | X.X |
| Min, Max | XXX.X, XXX.X | -XX.X, XX.X |

Table A4.15 Summary of Lab Changes – Hs-CRP

|  | ***Total (N=XX)*** | |
| --- | --- | --- |
|  | ***Absolute Result*** | ***Change From Baseline*** |
| **Hs-CRP (mg/L)** | | |
| **Baseline (Visit 1/Day 0)** | | |
| N | XX |  |
| Mean (SD) | X.X (XX.X) |  |
| Median | X.X |  |
| Min, Max | X.X, XX.X |  |
| **Visit 5/Day 14** | | |
| N | XX | XX |
| Mean (SD) | X.X (X.X) | -X.X (XX.X) |
| Median | X.X | -X.X |
| Min, Max | X.X, X.X | -XX.X, X.X |
| **Visit 6/Day 28** | | |
| N | XX | XX |
| Mean (SD) | X.X (X.X) | -X.X (XX.X) |
| Median | X.X | -X.X |
| Min, Max | X.X, X.X | -XX.X, X.X |

Table A4.16 Summary of Lab Changes – Ferritin

|  | ***Total (N=XX)*** | |
| --- | --- | --- |
|  | ***Absolute Result*** | ***Change From Baseline*** |
| **Ferritin (ng/mL)** | | |
| **Baseline (Visit 1/Day 0)** | | |
| N | XX |  |
| Mean (SD) | XXX.X (XXX.X) |  |
| Median | XXX.X |  |
| Min, Max | XX.X, XXX.X |  |
| **Visit 5/Day 14** | | |
| N | XX | XX |
| Mean (SD) | XXX.X (XXX.X) | -XX.X (XX.X) |
| Median | XXX.X | -XX.X |
| Min, Max | XX.X, XXX.X | -XXX.X, XXX.X |
| **Visit 6/Day 28** | | |
| N | XX | XX |
| Mean (SD) | XXX.X (XXX.X) | -XX.X (XXX.X) |
| Median | XXX.X | -XX.X |
| Min, Max | XX.X, XXX.X | -XXX.X, X.X |

Table A4.17 Summary of Lab Changes – A1c

|  | ***Total (N=XX)*** | |
| --- | --- | --- |
|  | ***Absolute Result*** | ***Change From Baseline*** |
| **A1c (%)** | | |
| **Baseline (Visit 1/Day 0)** | | |
| N | XX |  |
| Mean (SD) | X.X (X.X) |  |
| Median | X.X |  |
| Min, Max | X.X, XX.X |  |
| **Visit 5/Day 14** | | |
| N | X | X |
| Mean (SD) | X.X (X.X) | -X.X (X.X) |
| Median | X.X | X.X |
| Min, Max | X.X, X.X | -X.X, X.X |
| **Visit 6/Day 28** | | |
| N | X | X |
| Mean (SD) | X.X (X.X) | -X.X (X.X) |
| Median | X.X | -X.X |
| Min, Max | X.X, X.X | -X.X, X.X |

Table A4.18 Summary of Lab Changes – Fibrinogen

|  | ***Total (N=XX)*** | |
| --- | --- | --- |
|  | ***Absolute Result*** | ***Change From Baseline*** |
| **Fibrinogen (mg/dL)** | | |
| **Baseline (Visit 1/Day 0)** | | |
| N | XX |  |
| Mean (SD) | XXX.X (XX.X) |  |
| Median | XXX.X |  |
| Min, Max | XXX.X, XXX.X |  |
| **Visit 5/Day 14** | | |
| N | XX | XX |
| Mean (SD) | XXX.X (XX.X) | -XX.X (XX.X) |
| Median | XXX.X | -XX.X |
| Min, Max | XXX.X, XXX.X | -XXX.X, XX.X |
| **Visit 6/Day 28** | | |
| N | XX | XX |
| Mean (SD) | XXX.X (XX.X) | -XX.X (XX.X) |
| Median | XXX.X | -XX.X |
| Min, Max | XXX.X, XXX.X | -XXX.X, XX.X |

Table A4.19 Summary of Lab Changes – D-dimer

|  | ***Total (N=XX)*** | |
| --- | --- | --- |
|  | ***Absolute Result*** | ***Change From Baseline*** |
| **D-dimer (mg/L FEU)** | | |
| **Baseline (Visit 1/Day 0)** | | |
| N | XX |  |
| Mean (SD) | X.X (X.X) |  |
| Median | X.X |  |
| Min, Max | X.X, X.X |  |
| **Visit 5/Day 14** | | |
| N | XX | XX |
| Mean (SD) | X.X (X.X) | X.X (X.X) |
| Median | X.X | X.X |
| Min, Max | X.X, X.X | -X.X, X.X |
| **Visit 6/Day 28** | | |
| N | XX | XX |
| Mean (SD) | X.X (X.X) | X.X (X.X) |
| Median | X.X | X.X |
| Min, Max | X.X, X.X | -X.X, X.X |

Table A4.20 Summary of Lab Changes – PT

|  | ***Total (N=XX)*** | |
| --- | --- | --- |
|  | ***Absolute Result*** | ***Change From Baseline*** |
| **PT (sec)** | | |
| **Baseline (Visit 1/Day 0)** | | |
| N | XX |  |
| Mean (SD) | XX.X (X.X) |  |
| Median | XX.X |  |
| Min, Max | X.X, XX.X |  |
| **Visit 5/Day 14** | | |
| N | XX | XX |
| Mean (SD) | XX.X (X.X) | X.X (X.X) |
| Median | XX.X | X.X |
| Min, Max | XX.X, XX.X | -X.X, X.X |
| **Visit 6/Day 28** | | |
| N | XX | XX |
| Mean (SD) | XX.X (X.X) | X.X (X.X) |
| Median | XX.X | X.X |
| Min, Max | XX.X, XX.X | -X.X, X.X |

Table A4.21 Summary of Lab Changes – INR

|  | ***Total (N=XX)*** | |
| --- | --- | --- |
|  | ***Absolute Result*** | ***Change From Baseline*** |
| **INR** | | |
| **Baseline (Visit 1/Day 0)** | | |
| N | XX |  |
| Mean (SD) | X.X (X.X) |  |
| Median | X.X |  |
| Min, Max | X.X, X.X |  |
| **Visit 5/Day 14** | | |
| N | XX | XX |
| Mean (SD) | X.X (X.X) | X.X (X.X) |
| Median | X.X | X.X |
| Min, Max | X.X, X.X | -X.X, X.X |
| **Visit 6/Day 28** | | |
| N | XX | XX |
| Mean (SD) | X.X (X.X) | X.X (X.X) |
| Median | X.X | X.X |
| Min, Max | X.X, X.X | -X.X, X.X |

Table A5.1 Summary of Vital Sign Changes – Systolic BP

|  | ***Total (N=XX)*** | |
| --- | --- | --- |
|  | ***Absolute Result*** | ***Change From Baseline*** |
| **Systolic BP (mmHg)** | | |
| **Baseline (Visit 1/Day 0)** | | |
| N | XX |  |
| Mean (SD) | XXX.X (XX.X) |  |
| Median | XXX.X |  |
| Min, Max | XXX.X, XXX.X |  |
| **Visit 2/Day 2** | | |
| N | XX | XX |
| Mean (SD) | XXX.X (XX.X) | X.X (XX.X) |
| Median | XXX.X | -X.X |
| Min, Max | XX.X, XXX.X | -XX.X, XX.X |
| **Visit 3/Day 4** | | |
| N | XX | XX |
| Mean (SD) | XXX.X (XX.X) | -X.X (XX.X) |
| Median | XXX.X | -X.X |
| Min, Max | XXX.X, XXX.X | -XX.X, XX.X |
| **Visit 4/Day 6** | | |
| N | XX | XX |
| Mean (SD) | XXX.X (XX.X) | X.X (XX.X) |
| Median | XXX.X | X.X |
| Min, Max | XXX.X, XXX.X | -XX.X, XX.X |
| **Visit 5/Day 14** | | |
| N | XX | XX |
| Mean (SD) | XXX.X (XX.X) | X.X (XX.X) |
| Median | XXX.X | -X.X |
| Min, Max | XXX.X, XXX.X | -XX.X, XX.X |
| **Visit 6/Day 28** | | |
| N | XX | XX |
| Mean (SD) | XXX.X (XX.X) | X.X (XX.X) |
| Median | XXX.X | X.X |
| Min, Max | XXX.X, XXX.X | -XX.X, XX.X |

Table A5.2 Summary of Vital Sign Changes – Diastolic BP

|  | ***Total (N=XX)*** | |
| --- | --- | --- |
|  | ***Absolute Result*** | ***Change From Baseline*** |
| **Diastolic BP (mmHg)** | | |
| **Baseline (Visit 1/Day 0)** | | |
| N | XX |  |
| Mean (SD) | XX.X (XX.X) |  |
| Median | XX.X |  |
| Min, Max | XX.X, XXX.X |  |
| **Visit 2/Day 2** | | |
| N | XX | XX |
| Mean (SD) | XX.X (XX.X) | -X.X (X.X) |
| Median | XX.X | -X.X |
| Min, Max | XX.X, XXX.X | -XX.X, XX.X |
| **Visit 3/Day 4** | | |
| N | XX | XX |
| Mean (SD) | XX.X (XX.X) | -X.X (X.X) |
| Median | XX.X | -X.X |
| Min, Max | XX.X, XXX.X | -XX.X, XX.X |
| **Visit 4/Day 6** | | |
| N | XX | XX |
| Mean (SD) | XX.X (XX.X) | -X.X (X.X) |
| Median | XX.X | -X.X |
| Min, Max | XX.X, XXX.X | -XX.X, XX.X |
| **Visit 5/Day 14** | | |
| N | XX | XX |
| Mean (SD) | XX.X (XX.X) | -X.X (X.X) |
| Median | XX.X | -X.X |
| Min, Max | XX.X, XXX.X | -XX.X, XX.X |
| **Visit 6/Day 28** | | |
| N | XX | XX |
| Mean (SD) | XX.X (XX.X) | X.X (X.X) |
| Median | XX.X | X.X |
| Min, Max | XX.X, XXX.X | -XX.X, XX.X |

Table A5.3 Summary of Vital Sign Changes – Temperature

|  | ***Total (N=XX)*** | |
| --- | --- | --- |
|  | ***Absolute Result*** | ***Change From Baseline*** |
| **Temperature (F)** | | |
| **Baseline (Visit 1/Day 0)** | | |
| N | XX |  |
| Mean (SD) | XX.X (X.X) |  |
| Median | XX.X |  |
| Min, Max | XX.X, XX.X |  |
| **Visit 2/Day 2** | | |
| N | XX | XX |
| Mean (SD) | XX.X (X.X) | -X.X (X.X) |
| Median | XX.X | -X.X |
| Min, Max | XX.X, XX.X | -X.X, X.X |
| **Visit 3/Day 4** | | |
| N | XX | XX |
| Mean (SD) | XX.X (X.X) | -X.X (X.X) |
| Median | XX.X | -X.X |
| Min, Max | XX.X, XXX.X | -X.X, X.X |
| **Visit 4/Day 6** | | |
| N | XX | XX |
| Mean (SD) | XX.X (X.X) | -X.X (X.X) |
| Median | XX.X | X.X |
| Min, Max | XX.X, XXX.X | -X.X, X.X |
| **Visit 5/Day 14** | | |
| N | XX | XX |
| Mean (SD) | XX.X (X.X) | -X.X (X.X) |
| Median | XX.X | -X.X |
| Min, Max | XX.X, XX.X | -X.X, X.X |
| **Visit 6/Day 28** | | |
| N | XX | XX |
| Mean (SD) | XX.X (X.X) | -X.X (X.X) |
| Median | XX.X | -X.X |
| Min, Max | XX.X, XX.X | -X.X, X.X |

Table A5.4 Summary of Vital Sign Changes – Heart Rate

|  | ***Total (N=XX)*** | |
| --- | --- | --- |
|  | ***Absolute Result*** | ***Change From Baseline*** |
| **Heart Rate (bpm)** | | |
| **Baseline (Visit 1/Day 0)** | | |
| N | XX |  |
| Mean (SD) | XX.X (XX.X) |  |
| Median | XX.X |  |
| Min, Max | XX.X, XXX.X |  |
| **Visit 2/Day 2** | | |
| N | XX | XX |
| Mean (SD) | XX.X (XX.X) | -X.X (XX.X) |
| Median | XX.X | -X.X |
| Min, Max | XX.X, XXX.X | -XX.X, XX.X |
| **Visit 3/Day 4** | | |
| N | XX | XX |
| Mean (SD) | XX.X (XX.X) | -X.X (XX.X) |
| Median | XX.X | X.X |
| Min, Max | XX.X, XXX.X | -XX.X, XX.X |
| **Visit 4/Day 6** | | |
| N | XX | XX |
| Mean (SD) | XX.X (XX.X) | X.X (XX.X) |
| Median | XX.X | X.X |
| Min, Max | XX.X, XXX.X | -XX.X, XX.X |
| **Visit 5/Day 14** | | |
| N | XX | XX |
| Mean (SD) | XX.X (XX.X) | -X.X (XX.X) |
| Median | XX.X | -X.X |
| Min, Max | XX.X, XX.X | -XX.X, XX.X |
| **Visit 6/Day 28** | | |
| N | XX | XX |
| Mean (SD) | XX.X (X.X) | -X.X (XX.X) |
| Median | XX.X | -X.X |
| Min, Max | XX.X, XX.X | -XX.X, XX.X |

Table A5.5 Summary of Vital Sign Changes – Respiration Rate

|  | ***Total (N=XX)*** | |
| --- | --- | --- |
|  | ***Absolute Result*** | ***Change From Baseline*** |
| **Respiration Rate (bpm)** | | |
| **Baseline (Visit 1/Day 0)** | | |
| N | XX |  |
| Mean (SD) | XX.X (X.X) |  |
| Median | XX.X |  |
| Min, Max | XX.X, XX.X |  |
| **Visit 2/Day 2** | | |
| N | XX | XX |
| Mean (SD) | XX.X (X.X) | -X.X (X.X) |
| Median | XX.X | X.X |
| Min, Max | XX.X, XX.X | -X.X, X.X |
| **Visit 3/Day 4** | | |
| N | XX | XX |
| Mean (SD) | XX.X (X.X) | -X.X (X.X) |
| Median | XX.X | X.X |
| Min, Max | XX.X, XX.X | -X.X, X.X |
| **Visit 4/Day 6** | | |
| N | XX | XX |
| Mean (SD) | XX.X (X.X) | -X.X (X.X) |
| Median | XX.X | X.X |
| Min, Max | XX.X, XX.X | -X.X, X.X |
| **Visit 5/Day 14** | | |
| N | XX | XX |
| Mean (SD) | XX.X (X.X) | -X.X (X.X) |
| Median | XX.X | X.X |
| Min, Max | XX.X, XX.X | -X.X, X.X |
| **Visit 6/Day 28** | | |
| N | XX | XX |
| Mean (SD) | XX.X (X.X) | -X.X (X.X) |
| Median | XX.X | X.X |
| Min, Max | XX.X, XX.X | -X.X, X.X |

Table A5.6 Summary of Vital Sign Changes – Oxygen Saturation

|  | ***Total (N=XX)*** | |
| --- | --- | --- |
|  | ***Absolute Result*** | ***Change From Baseline*** |
| **Oxygen Saturation (%)** | | |
| **Baseline (Visit 1/Day 0)** | | |
| N | XX |  |
| Mean (SD) | XX.X (X.X) |  |
| Median | XX.X |  |
| Min, Max | XX.X, XXX.X |  |
| **Visit 2/Day 2** | | |
| N | XX | XX |
| Mean (SD) | XX.X (X.X) | X.X (X.X) |
| Median | XX.X | X.X |
| Min, Max | XX.X, XXX.X | -X.X, X.X |
| **Visit 3/Day 4** | | |
| N | XX | XX |
| Mean (SD) | XX.X (X.X) | X.X (X.X) |
| Median | XX.X | X.X |
| Min, Max | XX.X, XXX.X | -X.X, X.X |
| **Visit 4/Day 6** | | |
| N | XX | XX |
| Mean (SD) | XX.X (X.X) | -X.X (X.X) |
| Median | XX.X | X.X |
| Min, Max | XX.X, XXX.X | -XX.X, X.X |
| **Visit 5/Day 14** | | |
| N | XX | XX |
| Mean (SD) | XX.X (X.X) | X.X (X.X) |
| Median | XX.X | X.X |
| Min, Max | XX.X, XXX.X | -X.X, X.X |
| **Visit 6/Day 28** | | |
| N | XX | XX |
| Mean (SD) | XX.X (X.X) | X.X (X.X) |
| Median | XX.X | X.X |
| Min, Max | XX.X, XXX.X | -X.X, X.X |

Table A5.7 Summary of Vital Sign Changes – Weight

|  | ***Total (N=XX)*** | |
| --- | --- | --- |
|  | ***Absolute Result*** | ***Change From Baseline*** |
| **Weight (kg)** | | |
| **Baseline (Visit 1/Day 0)** | | |
| N | XX |  |
| Mean (SD) | XXX.X (XX.X) |  |
| Median | XXX.X |  |
| Min, Max | XX.X, XXX.X |  |
| **Visit 2/Day 2** | | |
| N | XX | XX |
| Mean (SD) | XXX.X (XX.X) | X.X (XX.X) |
| Median | XX.X | -X.X |
| Min, Max | XX.X, XXX.X | -XX.X, XX.X |
| **Visit 3/Day 4** | | |
| N | XX | XX |
| Mean (SD) | XXX.X (XX.X) | XX.X (XX.X) |
| Median | XXX.X | X.X |
| Min, Max | XX.X, XXX.X | -XX.X, XXX.X |
| **Visit 4/Day 6** | | |
| N | XX | XX |
| Mean (SD) | XXX.X (XX.X) | -X.X (XX.X) |
| Median | XX.X | X.X |
| Min, Max | XX.X, XXX.X | -XXX.X, XXX.X |
| **Visit 5/Day 14** | | |
| N | XX | XX |
| Mean (SD) | XXX.X (XX.X) | X.X (XX.X) |
| Median | XX.X | X.X |
| Min, Max | XX.X, XXX.X | -XXX.X, XXX.X |
| **Visit 6/Day 28** | | |
| N | XX | XX |
| Mean (SD) | XXX.X (XX.X) | -XX.X (XX.X) |
| Median | XX.X | X.X |
| Min, Max | XX.X, XXX.X | -XXX.X, XXX.X |
